# Transmission dynamics and forecasts of the COVID-19 pandemic in Mexico, March 20-November 11, 2020

**DOI:** 10.1101/2021.01.11.21249561

**Authors:** Amna Tariq, Juan M. Banda, Pavel Skums, Sushma Dahal, Carlos Castillo-Garsow, Baltazar Espinoza, Noel G. Brizuela, Roberto A. Saenz, Alexander Kirpich, Ruiyan Luo, Anuj Srivastava, Humberto Gutierrez, Nestor Garcia Chan, Ana I. Bento, Maria-Eugenia Jimenez-Corona, Gerardo Chowell

## Abstract

Mexico has experienced one of the highest COVID-19 death rates in the world. A delayed response towards implementation of social distancing interventions until late March 2020 and a phased reopening of the country in June 2020 has facilitated sustained disease transmission in the region. Here, we systematically generate and compare 30-day ahead forecasts using previously validated growth models based on mortality trends from the Institute for Health Metrics and Evaluation for Mexico and Mexico City in near real-time. Moreover, we estimate reproduction numbers for SARS-CoV-2 based on methods that rely on genomic data as well as case incidence data. Subsequently, functional data analysis techniques are utilized to analyze the shapes of COVID-19 growth rate curves at the state level to characterize the spatial-temporal transmission patterns. The early estimates of reproduction number for Mexico were estimated between R∼1.1-from genomic and case incidence data. Moreover, the mean estimate of R has fluctuated ∼1.0 from late July till end of September 2020. The spatial analysis characterizes the state-level dynamics of COVID-19 into four groups with distinct epidemic trajectories. We found that the sequential mortality forecasts from the GLM and Richards model predict downward trends in the number of deaths for all thirteen forecasts periods for Mexico and Mexico City. The sub-epidemic and IHME models predict more realistic stable trajectory of COVID-19 mortality trends for the last three forecast periods (09/21-10/21 - 09/28-10/27) for Mexico and Mexico City. Our findings support the view that phenomenological models are useful tools for short-term epidemic forecasting albeit forecasts need to be interpreted with caution given the dynamic implementation and lifting of social distancing measures.

## Introduction

The ongoing COVID-19 (coronavirus disease 2019) pandemic is the most important global health challenge since the 1918 influenza pandemic [1]. This calls for scientists, health professionals and policy makers to collaboratively address the challenges posed by this deadly infectious disease. The causative SARS-CoV-2 (severe acute respiratory syndrome virus 2) is a novel, unusually complex and highly transmissible virus that spreads via respiratory droplets and aerosols [2, 3]. It presents a clinical spectrum that ranges from asymptomatic individuals to conditions that require the use of mechanical ventilation to multiorgan failure and septic shock leading to death [2]. The ongoing COVID-19 pandemic has not only exerted significant morbidity but also excruciating mortality burden with more than 79.2 million cases and 1.7 million deaths reported worldwide as of December 29, 2020 [4]. Approximately 27 countries globally including 9 countries in the Americas have reported more than 10,000 deaths attributable to SARS-CoV-2 as of December 29, 2020 despite the implementation of social distancing policies to limit the death toll [5]. In comparison, a total of 774 deaths were reported during the 2003 SARS multi-country epidemic and 858 deaths were reported during the 2012 MERS epidemic in Saudi Arabia [6, 7].

Determining the best containment strategies for COVID-19 pandemic is a highly active research area [3]. While multiple vaccines against the novel coronavirus have begun to roll out, many scientific uncertainties exist that will dictate how vaccination campaigns will affect the course of the pandemic. For instance, it is still unclear if the vaccine will prevent the transmission of SARS-CoV-2 or just protect against more severe disease outcomes and death [8, 9]. In these circumstances non-pharmaceutical interventions remain the most promising policy levers to reduce the virus transmission [10]. The epidemiological and mathematical models can help quantify the effects of these non-pharmaceutical interventions such as wearing facemasks and social distancing mandates to contain the spread of the virus [11]. However, recent studies have demonstrated that indicators such as population density, poverty, over-crowding and inappropriate work place conditions hinder the social distancing interventions propagating the unmitigated spread of the virus, especially in developing countries [12, 13]. Moreover, the differential mortality trends are also influenced by the disparate disease burden driven by the socioeconomic gradients with the poorest areas showing highest preventable mortality rates [14].

Mexico, exhibiting one of the highest COVID-19 mortality impact in the world thus far [15], is a highly populated country [16] with ∼42% of the population living in poverty (defined as the state if a person or group of people lack a specified amount of money or material possessions) [17] and ∼60% of the population work in the informal sector [18]. In this context, Mexico ranks fourth in the world in terms of the number of COVID-19 deaths, a tally surpassed only by the USA, Brazil and India [19], has reported one of highest death tolls among healthcare workers (∼2500 deaths) [20], and has conducted the lowest number of COVID-19 tests per capita [21].

The Mexican Ministry of Health identified three phases of contingency plan: viral importation, community transmission and epidemic to combat the COVID-19 pandemic in Mexico [22]. The pandemic was likely seeded by imported COVID-19 cases reported on February 28, 2020 [23, 24]. As the virus spread across the nation in phase one of the pandemic, some universities switched to virtual classes and some festivals and sporting events were postponed [25]. However, the government initially downplayed the impact of the virus and did not enforce strict social distancing measures [26]. This led to large gatherings at some social events such as concerts, festivals and soccer tournament amidst sustained disease transmission in the country [27]. A study conducted in Mexico estimated the early reproduction number for the first ten days of the epidemic between ∼2.9-4.9 [28]. However, the true impact of the pandemic was generally under-estimated by the Mexican government despite active virus transmission in the country [29].

As local clusters of disease started to appear in the community, phase 2 (community transmission) of the pandemic was declared on March 24, 2020 [30]. Authorities suspended all non-essential activities including closure of public and entertainment places and banned gatherings of more than 100 people [31]. This was followed by the declaration of national emergency on March 30, 2020. The new measures to fight the virus under the national emergency included extending the suspension of non-essential activities and a reduction the number of people who can gather to fifty [32]. However, as the virus paved its way across the country ravaging through the poor and rural communities, the government urged the public to comply with the stay-at-home orders [33-35]. These preventive orders from the government were met with mixed reactions from people belonging to different socio-economic sectors of the community [36]. Moreover, transportation restrictions to and from the regions most affected by COVID-19 were not implemented until April 16, 2020 [37]. Shortly after, on April 21, 2020, Mexico announced phase 3 of the contingency (epidemic phase) as wide-spread community transmission intensified [38].

With lockdowns and other restrictions in place, Mexican officials shared model output [39] predicting that COVID-19 case counts would peak in early May and that the epidemic was expected to end before July 2020 [40]. Despite notorious disagreement between surveillance data and government forecasts, these model predictions continued to be cited by official and independent sources [41, 42]. The extent to which these overly optimistic predictions skewed the plans and budgets of private and public institutions remains unknown. Under the official narrative that the pandemic would soon be over, Mexico planned a gradual phased re-opening of its economy in early June 2020, as the “new normal” phase [43].

In Mexico, reopening of the economic activities started on June 1 under a four color traffic light monitoring system to alert the residents of the epidemiological risks of COVID-19 based on the level of severity of the pandemic in each state, on a weekly basis [44, 45]. As of December 29, 2020 Mexico exhibits high estimates of cumulative COVID-19 cases and deaths; 1,401,529 and 123,845 respectively [15]. Given the high transmission potential of the virus and limited application of tests in the country, testing only 24.54 people for every 1000 people (as of December 28, 2020) [21], estimates of the effective reproduction number from the case incidence data and near real-time epidemic projections using mortality data could prove to be highly beneficial to understand the epidemic trajectory of COVID-19. It may also be useful to assess the effect of intervention strategies such as the stay-at-home orders and mobility patterns on the epidemic curve and understand the different spatiotemporal dynamics of the virus.

In order to investigate the transmission dynamics of the unfolding COVID-19 epidemic in Mexico, we analyze the case data by date of symptoms onset and death data by date of reporting utilizing mathematical models that are useful to characterize the empirical patterns of epidemics [46, 47]. We estimate the effective reproduction number of SARS-CoV-2 in Mexico to understand the transmission dynamics of the virus and examine the mobility trends in relation to the curve of the number of COVID-19 deaths. Moreover, we employ statistical methods from functional data analysis to study the shapes of the COVID-19 growth rate curves at the state level. This helps us characterize the spatial-temporal dynamics of the pandemic based on the shape features of these curves. Lastly, twitter data demonstrating the frequency of tweets indicating stay-at-home-order is analyzed in relation to the COVID-19 case counts at the national level.

## Methods

### Data

Five sources of data are analyzed in this manuscript. A brief description of the datasets and their sources are described below.

#### (i) IHME data for short term forecasts

We utilized the openly published smoothed trend in daily COVID-19 reported deaths from the Institute of Health Metrics and Evaluation (IHME) for (i) Mexico (country) and (ii) Mexico City (capital of Mexico) as of October 9, 2020 to generate the forecasts [48]. IHME smoothed data estimates (current projection scenario) were utilized as they were corrected for the irregularities in the daily death data reporting, by averaging model results over the last seven days. As this was our source of data for prediction modeling, it was chosen for its consistent updates. The statistical procedure of spline regressions obtained from MR-BRT (“meta-regression—Bayesian, regularized, trimmed”) smooths the trend in COVID-19 reported deaths as described in ref [10].

This data are publicly available from the IHME COVID-19 estimates downloads page [48]. For this analysis, deaths as reported by the IHME model (current projection scenario as described ahead) on November 11, 2020 are used as a proxy for actual reported deaths attributed to COVID-19.

#### (ii) Apple mobility trends data

Mobility data for Mexico published publicly by Apple’s mobility trends reports was retrieved as of December 5, 2020 [49]. This aggregated and anonymized data is updated daily and includes the relative volume of directions requests per country compared to a baseline volume on January 13, 2020. Apple has released the data for the three modes of human mobility: driving, walking and public transit. The mobility measures are normalized in the range 0-100 for each country at the beginning of the series, so trends are relative to this baseline.

#### (iii) Case incidence and genomic data for estimating reproduction number

In order to estimate the reproduction number, we use two different data sources. For estimating the early reproduction number from the genomic data, 111 SARS-CoV-2 genome samples were obtained from the “global initiative on sharing avian influenza data” (GISAID) repository between February 27-May 29, 2020 [50]. For estimating the reproduction number from the case incidence data (early reproduction number and the instantaneous reproduction number), we utilized publicly available time series of laboratory-confirmed cases by dates of symptoms onset which were obtained from the Mexican Ministry of Health as of December 5, 2020 [15].

#### (iv) Case incidence data for Spatial analysis

We recovered daily case incidence data for all 32 states of Mexico from March 20 to December 5 from the Ministry of Health Mexico, as of December 5, 2020 [15].

#### (v) Twitter data for twitter analysis

For the twitter data analysis, we retrieved data from the publicly available twitter data set of COVID-19 chatter from March 12 to November 11, 2020 [51].

### Modeling framework for forecast generation

We harness three dynamic phenomenological growth models previously applied to multiple infectious diseases (e.g., SARS, foot and mouth disease, Ebola [52, 53] and the current COVID-19 outbreak [54, 55]) for mortality modeling and short-term forecasting in Mexico and Mexico City. These models include the simple scalar differential equation models such as the generalized logistic growth model [53] and the Richards growth model [56]. We also utilize the sub-epidemic wave model [52] which accommodates complex epidemic trajectories by assembling the contribution of multiple overlapping sub-epidemic waves. The mortality forecasts obtained from these mathematical models can provide valuable insights on the disease transmission mechanisms, the efficacy of intervention strategies and help evaluate optimal resource allocation procedures to inform public health policies. The COVID-19 mortality forecasts for Mexico and Mexico City generated by IHME (current projections scenario) are used as a benchmark model. The description of these models is provided in the supplemental file.

Cumulative mortality forecasts obtained from our phenomenological growth models are compared with the total mean smoothed death data estimates retrieved from the IHME reference scenario and two IHME counterfactual scenarios. The IHME reference scenario depicts the “current projection”, which assumes that the social distancing measures are re-imposed for six weeks whenever daily deaths reach eight per million. The second scenario “mandates easing” implies what would happen if the government continues to ease social distancing measures without re-imposition. Lastly, the third scenario, “universal masks” accounts for universal facemask wearing, that reflects 95% facemask usage in public and social distancing mandates reimposed at 8 deaths per million. Detailed description of these modeling scenarios and their assumptions is explained in ref. [10]. Moreover, the total mean smoothed death data estimates reported by IHME reference scenario as of November 11, 2020 are considered as a proxy for actual death count for each forecasting period.

### Model calibration and forecasting approach

We conducted 30-day ahead short-term forecasts utilizing thirteen data sets spanned over a period of four months (July 4-October 9, 2020) (Table 1). Each forecast was fitted to the daily death counts from the IHME smoothed data estimates reported between March 20-September 27, 2020 for (i) Mexico and (ii) Mexico City. The first model calibration process relies on fifteen weeks of data, from March 20-July 4, 2020. Sequentially models are recalibrated each week with the most up-to-date data, meaning the length of the calibration period increases by one week up to August 2, 2020. However, owing to irregular publishing of data estimates by the IHME, the length of calibration period increased by 2 weeks after August 2, 2020. This was followed by a one week increase from August 17-September 27, 2020 as the data estimates were again published every week.

**Table 1:**
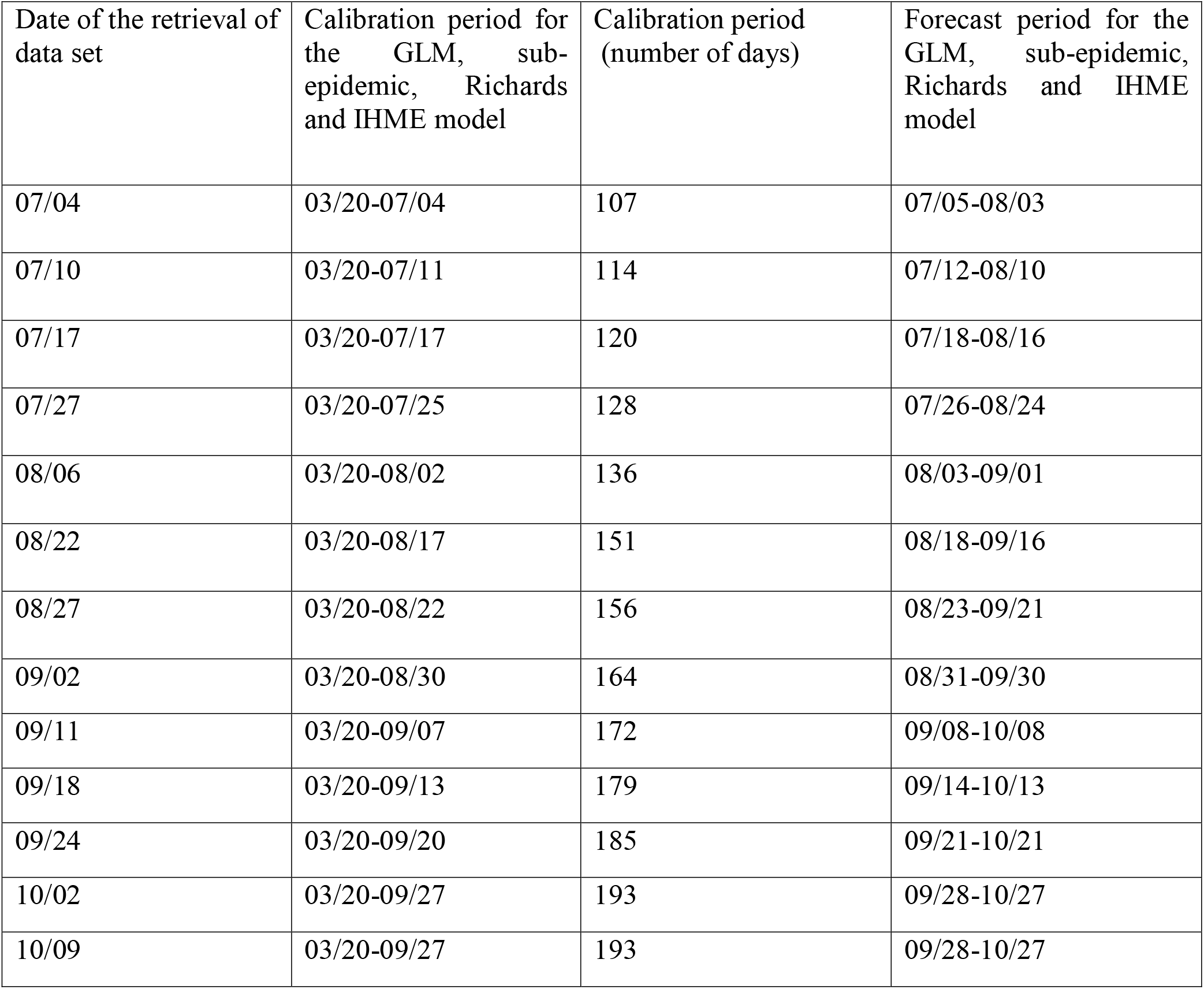
Characteristics of the data sets used for the sequential calibration and forecasting of the COVID-19 epidemic in Mexico and Mexico City (2020).

The 30-day ahead shot-term forecasts generated by calibrating our three phenomenological growth models with the IHME smoothed death data estimates are compared with the forecasts generated by the IHME reference scenario for the same calibration and forecasting periods.

For each of the three models; GLM, Richards growth model and the sub-epidemic wave model, we estimate the best fit solution for each model using non-linear least square fitting procedure [57]. This process minimizes the sum of squared errors between the model fit, 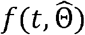 and the smoothed data estimates, *y*_*t*_ and yields the best set of parameter estimates Θ = (θ_1_, θ_2_, …, θ_m_). The parameters 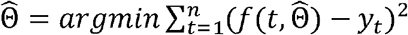 define the best fit model *f*(*t, Θ*). Here 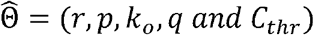 corresponds to the set of parameters of the sub-epidemic model, 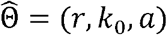 corresponds to set of parameters of the Richards model and 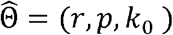 corresponds to the set of parameters of the GLM model [58]. For the GLM and sub-epidemic wave model, we provide initial best guesses of the parameter estimates. However, for the Richards growth model we initialize the parameters for the nonlinear least squares’ method [57] over a wide range of plausible parameters from a uniform distribution using Latin hypercube sampling [59]. This allows us to test the uniqueness of the best fit model. Moreover, the initial conditions are set at the first data point for each of the three models [58]. Uncertainty bounds around the best-fit solution are generated using parametric bootstrap approach with replacement of data assuming a Poisson error structure for the GLM and sub-epidemic model. A negative binomial error structure was used to generate the uncertainty bounds of the Richards growth model; where we assume the mean to be three times the variance based on the noise in the data. Detailed description of this method is provided in ref [58].

Each of the M best-fit parameter sets are used to construct the 95% confidence intervals for each parameter by refitting the models to each of the M = 300 datasets generated by the bootstrap approach during the calibration phase. Further, each M best fit model solution is used to generate m= 30 additional simulations with Poisson error structure for GLM and sub-epidemic model and negative binomial error structure for Richards model extended through a 30-day forecasting period. For the forecasting period, we construct the 95% prediction intervals with these 9000 (M × m) curves. Detailed description of the methods of parameter estimation can be found in references [58, 60, 61]

### Performance metrics

We utilized the following four performance metrics to assess the quality of our model fit and the 30-day ahead short term forecasts: the mean absolute error (MAE) [62], the mean squared error (MSE) [63], the coverage of the 95% prediction intervals [63], and the mean interval score (MIS) [63] for each of the three models: GLM, Richards model and the sub-epidemic model. For calibration performance, we compare the model fit to the observed smoothed death data estimates fitted to the model, whereas for the performance of forecasts, we compare our forecasts with the smoothed death data estimates (current projections scenario) reported on November 11, 2020 for the time period of the forecast.

The mean squared error (MSE) and the mean absolute error (MAE) assess the average deviations of the model fit to the observed death data. The mean absolute error (MAE) is given by:

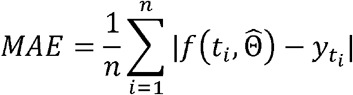

The mean squared error (MSE) is given by:

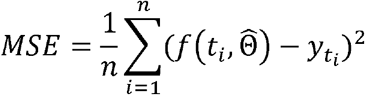

where 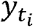 is the time series of reported smoothed death estimates, *t*_*i*_ is the time stamp and 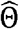 is the set of model parameters. For the calibration period, *n* equals the number of data points used for calibration, and for the forecasting period, *n =* 30 for the 30-day ahead short-term forecast.

Moreover, in order to assess the model uncertainty and performance of prediction interval, we use the 95% PI and MIS. The prediction coverage is defined as the proportion of observations that fall within 95% prediction interval and is calculated as:

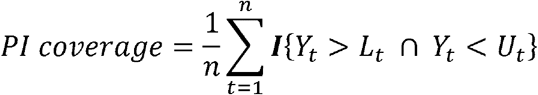

where *Y*_*t*_ are the smoothed death data estimates, *L*_*t*_ and *U*_*t*_ are the lower and upper bounds of the 95% prediction intervals, respectively, *n* is the length of the period, and I is an indicator variable that equals 1 if value of *Y*_*t*_ is in the specified interval and 0 otherwise

The mean interval score addresses the width of the prediction interval as well as the coverage. The mean interval score (MIS) is given by:

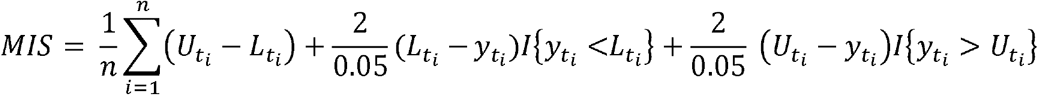

In this equation *L*_*t*_, *U*_*t*_, *n* and I are as specified above for PI coverage. Therefore, if the PI coverage is 1, the MIS is the average width of the interval across each time point. For two models that have an equivalent PI coverage, a lower value of MIS indicates narrower intervals [63].

### Mobility data analysis

In order to analyze the time series data for Mexico from March 20-December 5, 2020 for three modes of mobility; driving, walking and public transport, we utilize the R code developed by Kieran Healy [64]. We analyze the mobility trends to look for any pattern in sync with the mortality curve of COVID-19. The time series for mobility requests is decomposed into trends, weekly and remainder components. The trend is a locally weighted regression fitted to the data and remainder is any residual left over on any given day after the underlying trend and normal daily fluctuations have been accounted for.

### Reproduction number

We estimate the reproduction number, *R*_*t*_, for the early ascending phase of the COVID-19 epidemic in Mexico and the instantaneous reproduction number *R*_*t*_ throughout the epidemic. Reproduction number, *R*_*t*_, is a crucial parameter that characterizes the average number of secondary cases generated by a primary case at calendar time *t* during the outbreak. This quantity is critical to identify the magnitude of public health interventions required to contain an epidemic [65-67]. Estimates of *R*_*t*_ indicate if widespread disease transmission continues (*R*_*t*_>1) or disease transmission declines (*R*_*t*_<1). Therefore, in order to contain an outbreak, it is vital to maintain *R*_*t*_<1.

### Estimating the reproduction number, *R*_*t*_, from case incidence using generalized growth model (GGM)

We estimate the reproduction number by calibrating the GGM (as described in the supplemental file) to the early growth phase of the epidemic (February 27-May 29, 2020) [68]. The generation interval of SARS-CoV-2 is modeled assuming gamma distribution with a mean of 5.2 days and a standard deviation of 1.72 days [69]. We estimate the growth rate parameter *r*, and the deceleration of growth parameter *p*, as described in the supplemental file. The GGM model is used to simulate the progression of local incidence cases *I*_*i*_ at calendar time *t*_*i*_. This is followed by the application of the discretized probability distribution of the generation interval, denoted by *ρ*_*i*_, to the renewal equation to estimate the reproduction number at time *t*_*i*_ [70-72]:

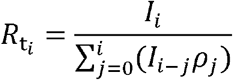

The numerator represents the total new cases *I*_*i*_, and the denominator represents the total number of cases that contribute (as primary cases) to generating the new cases *I*_*i*_ (as secondary cases) at time *t*_*i*_. This way, *R*_*t*_, represents the average number of secondary cases generated by a single case at calendar time *t*. The uncertainty bounds around the curve of *R*_*t*_ are derived directly from the uncertainty associated with the parameter estimates (*r, p*) obtained from the GGM. We estimate *R*_*t*_ for 300 simulated curves assuming a negative binomial error structure [58].

### Instantaneous reproduction number *R*_*t*_, using the Cori method

The instantaneous *R*_*t*_ is estimated by the ratio of number of new infections generated at calendar time *t*(*I*_*t*_), to the total infectiousness of infected individuals at time *t* given by 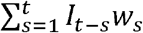 [73,74]. Hence *R*_*t*_ can be written as:

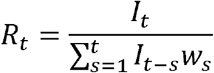

In this equation, *I*_*t*_ is the number of new infections on day *t* and *w*_*s*_ represents the infectivity function, which is the infectivity profile of the infected individual. This is dependent on the time since infection (*s)*, but is independent of the calendar time (*t*) [75, 76].

The term 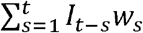 describes the sum of infection incidence up to time step *t* − 1, weighted by the infectivity function *w*_*s*_. The distribution of the generation time can be applied to approximate *w*_*s*_, however, since the time of infection is a rarely observed event, measuring the distribution of generation time becomes difficult [73]. Therefore, the time of symptom onset is usually used to estimate the distribution of serial interval (SI), which is defined as the time interval between the dates of symptom onset among two successive cases in a disease transmission chain [77].

The infectiousness of a case is a function of the time since infection, which is proportional to *w*_*s*_ if the timing of infection in the primary case is set as time zero of *w*_*s*_ and we assume that the generation interval equals the SI. The SI was assumed to follow a gamma distribution with a mean of 5.2 days and a standard deviation of 1.72 days [69]. Analytical estimates of *R*_*t*_ were obtained within a Bayesian framework using EpiEstim R package in R language [77]. *R*_*t*_ was estimated at weekly intervals. We reported the median and 95% credible interval (CrI).

### Estimating the reproduction number, *R*, from the genomic analysis

In order to estimate the reproduction number for the SARS-CoV-2 between February 27-May 29, 2020, from the genomic data, 111 SARS-CoV-2 genomes sampled from infected patients from Mexico and their sampling times were obtained from GISAID repository [50]. Short sequences and sequences with significant number of gaps and non-identified nucleotides were removed, yielding 83 high-quality sequences. For clustering, they were complemented by sequences from other geographical regions, down sampled to n=4325 representative sequences. We used the sequence subsample from Nextstrain (www.nextstrain.org) global analysis as of August 15, 2020. These sequences were aligned to the reference genome taken from the literature [78] using MUSCLE [79] and trimmed to the same length of 29772 bp. The maximum likelihood phylogeny has been constructed using RAxML[80]

The largest Mexican cluster that possibly corresponds to within-country transmissions has been identified using hierarchical clustering of sequences. The phylodynamics analysis of that cluster have been carried out using BEAST v1.10.4 [81]. We used strict molecular clock and the tree prior with exponential growth coalescent. Markov Chain Monte Carlo sampling has been run for 10,000,000 iterations, and the parameters were sampled every 1000 iterations. The exponential growth rate *f* estimated by BEAST was used to calculate the reproductive number *R*. For that, we utilized the standard assumption that SARS-CoV-2 generation intervals (times between infection and onward transmission) are gamma-distributed [82]. In that case *R* can be estimated as 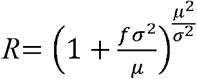 where *µ* and *σ* are the mean and standard deviation of that gamma distribution. Their values were taken from ref [69].

### Spatial analysis

For the shape analysis of incidence rate curves we followed ref. [83] to pre-process the daily cumulative COVID-19 case data at state level as follows:

a. Time differencing: If *f*_*i*_(*t*) denotes the given cumulative number of confirmed cases for state *i* on day *t* then per day growth rate at time *t* is given by *g*_*i*_ (*t*) = *f*_*i*_(*t*) − *f*_*i*_(*t* − 1).
b. Smoothing: We then smooth the normalized curves using smooth function in Matlab.
c. Rescaling: Rescaling of each curve is done by dividing each *g*_*i*_ by the total confirmed cases for a state *i* That is, compute *h*_*i*_ (*t*) = *g*_*i*_ (*t*) /*r*_*i*_ where *r* _*i*_ = ∑_*t*_*g*_*i*_ (*t*)

To identify the clusters by comparing the curves, we used a simple metric. For any two rate curves, *h*_*i*_ and *h*_*j*_, we compute the norm ||*h*_*i*_ −*h*_*j*_||, where the double bars denote the *L*^*2*^ norm of the difference function, i.e., 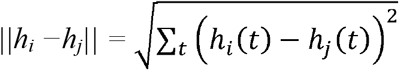.

This process is depicted in S17 Fig. To identify the clusters by comparing the curves, we used a simple metric. For any two rate curves, *h*_*i*_ and *h*_*j*_, we compute the norm ||*h*_*i*_*-h*_*j*_||, where the double bars denote the L^2^ norm of the difference function, i.e.,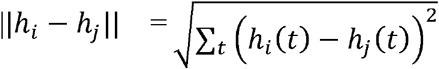. To perform clustering of 32 curves into smaller groups, we apply the dendrogram function in Matlab using the “ward” linkage as explained in ref [84]. The number of clusters is decided empirically based on the display of overall clustering results. After clustering the states into different groups, we derived average curve for each cluster after using a time wrapping algorithm as performed in refs [84, 85].

### Twitter data analysis

To observe any relationship between the COVID-19 cases by date of symptoms onset and the frequency of tweets indicating stay-at-home orders we used a public dataset of 698 million tweets of COVID-19 chatter [51]. The frequency of tweets indicating stay-at-home order is used to gauge the compliance of people with the orders of staying at home to avoid spread of the virus by maintaining social distance. Tweets indicate the magnitude of the people being pro-lockdown and depict how these numbers have dwindled over the course of the pandemic. To get to the plotted data, we removed all retweets and tweets not in the Spanish language. We also filtered by the following hashtags: #quedateencasa, and #trabajardesdecasa, which are two of the most used hashtags when users refer to the COVID-19 pandemic and their engagement with health measures. Lastly, we limited the tweets to the ones that originated from Mexico, via its 2-code country code: MX. A set of 521,359 unique tweets were gathered from March 12 to November 11, 2020. We then overlay the curve of tweets over the epi-curve in Mexico to observe any relation between the shape of the epidemic trajectory and the shape of curve for the frequency of tweets during the established time period. We also estimate the correlation coefficient between the cases and frequency of tweets.

## Results

As of November 11, 2020, Mexico has reported 105,656 deaths whereas Mexico City has reported 15,742 deaths as per the IHME smoothed death data estimates. Fig 1 (upper panel) shows the daily COVID-19 death curve in Mexico and Mexico City from March 20-November 11, 2020. The mobility trend for Mexico (Fig 1, lower panel) shows that the human mobility tracked in the form of walking, driving and public transportation declined from end of March to the beginning of June, corresponding to the implementation of social distancing interventions and the *Jornada Nacional de Sana Distancia* that was put in place between March 23-May 30, 2020 encompassing the suspension of non-essential activities in public, private and social sectors [86]. The driving and walking trend subsequently increased in June with the reopening of the non-essential services. Fig 1 (upper panel) shows that reopening of the country coincides with the highest levels of daily deaths. These remain at a high level for just over two months (June and July). Then, from mid-August, the number of deaths begins to fall, reaching a reduction of nearly 50% by mid-October. But at the end of October a new growth begins.

**Fig 1:**
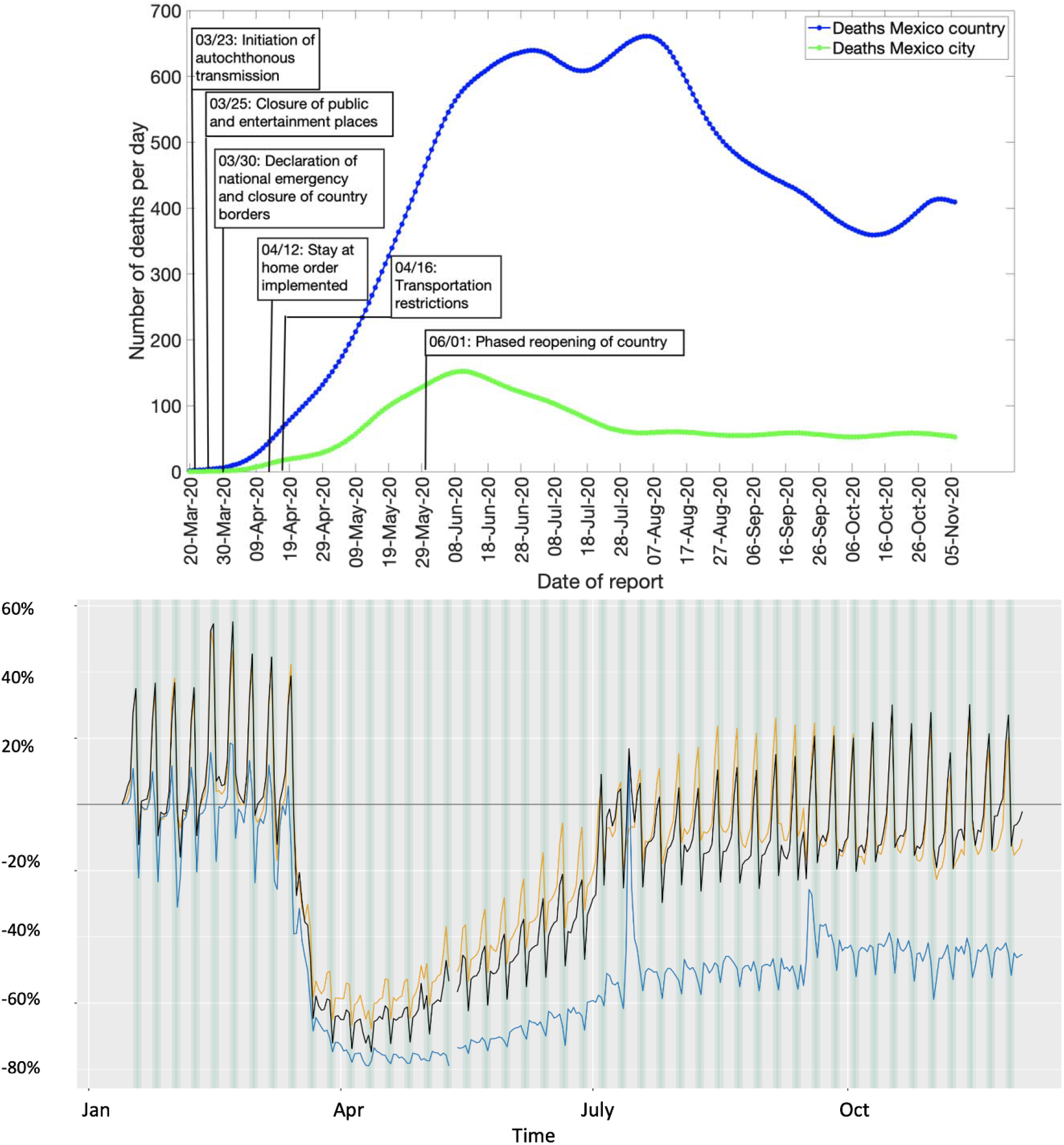
Upper panel: Epidemic curve for the COVID-19 deaths in Mexico and Mexico City from March 20-November 11, 2020. Blue line depicts the confirmed deaths in Mexico and green line depicts the confirmed deaths in Mexico City. Lower panel: The mobility trends for Mexico. Orange line shows the driving trend, blue line shows the transit trend and the black line shows the walking trend.

In the subsequent sections, we first present the results for the short-term forecasting, followed by the estimation of the reproduction numbers. Then we present the results of the spatial analysis and the twitter data analysis.

### Model calibration and forecasting performance

Here we compare the calibration and 30-day ahead forecasting performance of our three models: the GLM, Richards growth model and the sub-epidemic wave model between March 20-September 27, 2020 and July 5-October 27, 2020 respectively for (i) Mexico and (ii) Mexico City. We also compare the results of our cumulative mortality forecasts with the total mean smoothed death data estimates retrieved from the three IHME model scenarios (as explained in the methods section).

### Calibration performance

Across the thirteen sequential model calibration phases for Mexico over a period of seven months (March-September), as provided in Table S1 and Fig 2, the sub-epidemic model outperforms the GLM with lower RMSE estimates for the seven calibration phases 3/20-07/04, 3/20-7/17, 3/20-8/17, 3/20-08/22, 3/20-09/13, 3/20-09/20, 3/20-09/27. The GLM model outperforms the other two models for the remaining six calibration phases in terms of RMSE. The Richards model has substantially higher RMSE (between 10.2-24.9) across all thirteen calibration phases indicating a sub-optimal model fit. The sub-epidemic model also outperforms the other two models in terms of MAE, MIS and the 95% PI coverage. It has the lowest values for MIS and the highest 95% PI coverage for nine of the thirteen calibration phases (Table S1). Moreover, the sub-epidemic model has the lowest MAE for eleven calibration phases. The Richards model shows much higher MIS and lower 95% PI coverage compared to the GLM and sub-epidemic model, pointing towards a sub-optimal model fit.

**Fig 2:**
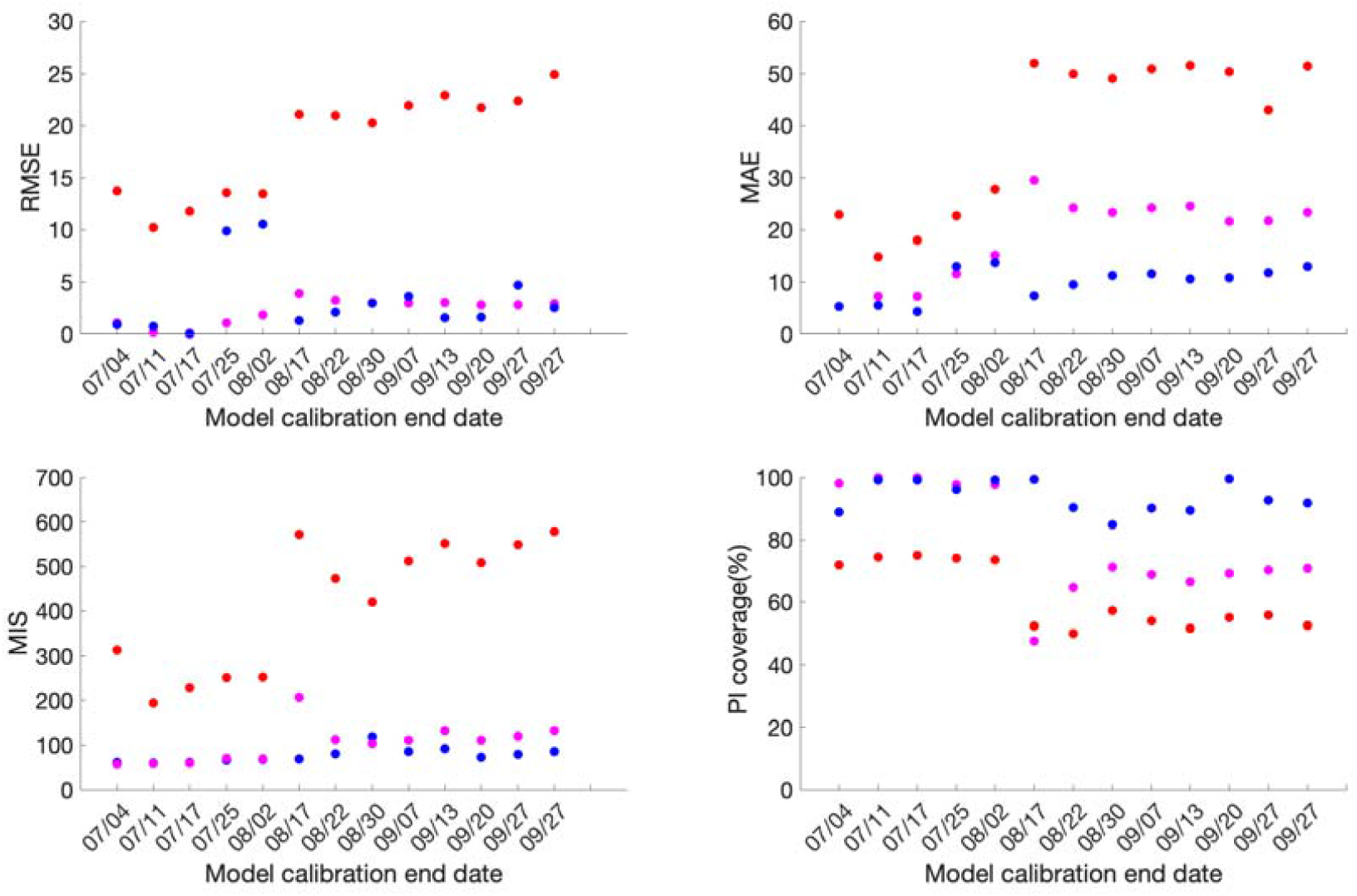
Calibration performance for each of the thirteen sequential calibration phases for GLM (magenta), Richards (red) and sub-epidemic (blue) model for Mexico. High 95% PI coverage and lower mean interval score (MIS), root mean square error (RMSE) and mean absolute error (MAE) indicate better performance.

For Mexico City, the sub-epidemic model outperforms the other two models in terms of all performance metrics. It has the lowest RMSE for eleven of the thirteen calibration phases followed by the GLM and Richards model. The MAE is also the lowest for the sub-epidemic model for all thirteen calibration phases, followed by the GLM and Richards growth model. Further, in terms of MIS, the sub-epidemic model outperforms the Richards and GLM model for nine calibration phases whereas the GLM model outperforms the other two models in four calibration phases (3/20-7/04, 3/20-7/11, 3/20-7/17, 3/20-8/02). The Richards model has much higher estimates for the MIS compared to the other two models indicating a sub-optimal model fit. The 95% PI across all thirteen calibration phases lies between 91.6-99.4% for the sub-epidemic model, followed by the Richards model (85.9-100%) and the GLM model (53.2-100%) (Table S2, Fig 3).

**Fig 3:**
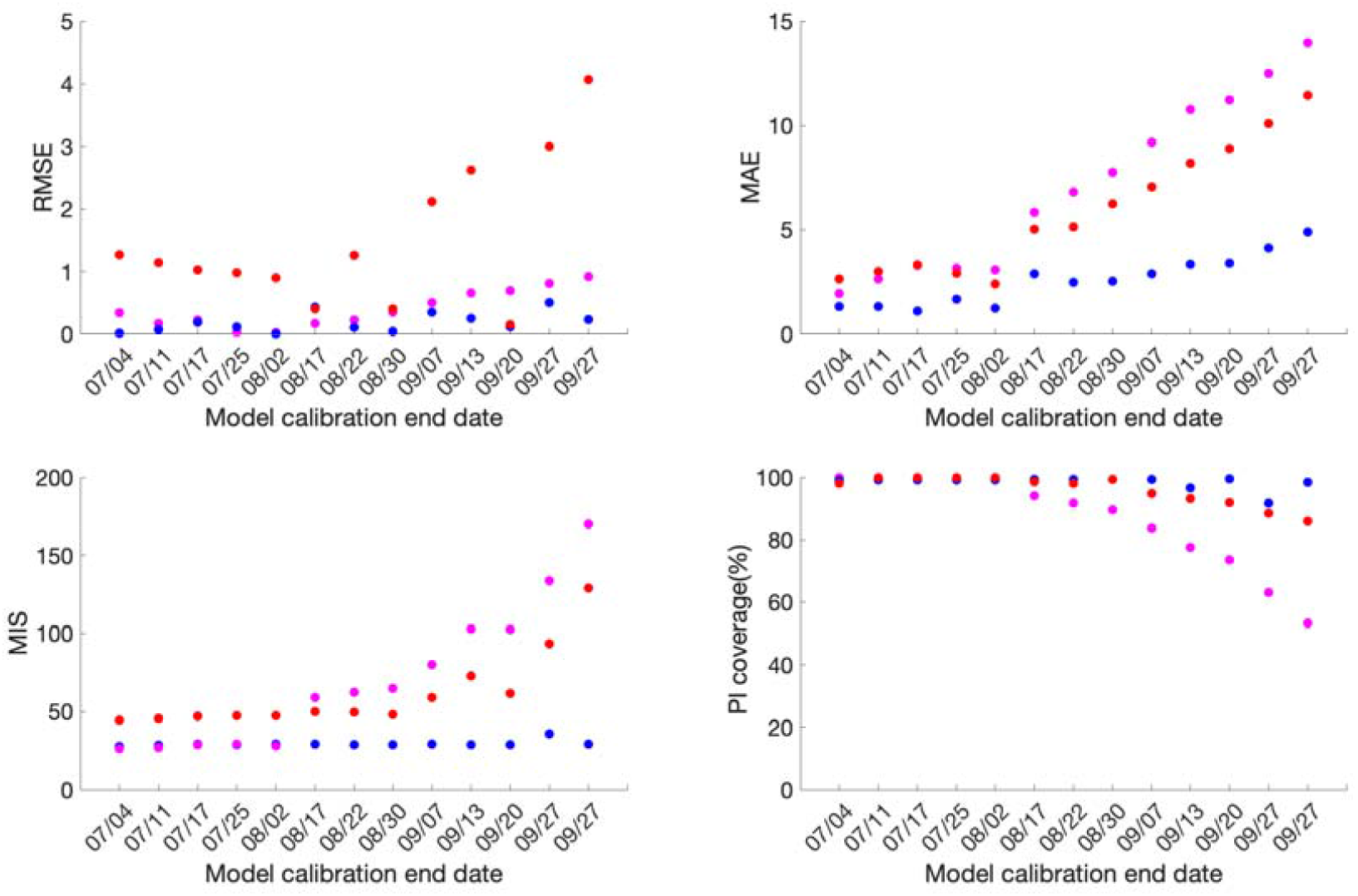
Calibration performance for each of the thirteen sequential calibration phases for GLM (magenta), Richards (red) and sub-epidemic (blue) model for Mexico City. High 95% PI coverage and lower mean interval score (MIS), root mean square error (RMSE) and mean absolute error (MAE) indicate better performance.

Over-all the goodness of fit metrics points toward the sub-epidemic model as the most appropriate model for the Mexico City and Mexico across all four-performance metrics except for the RMSE for Mexico, where the estimates of GLM model compete with the sub-epidemic model.

### Forecasting performance

For Mexico, the sub-epidemic model consistently outperforms the GLM and Richards growth model for ten out of the thirteen forecasting phases in terms of RMSE and MAE, eight forecasting phases in terms of MIS and nine forecasting phases in terms of the 95% PI coverage. This is followed by the GLM and the Richards growth model (Fig 4, Table S4).

**Fig 4:**
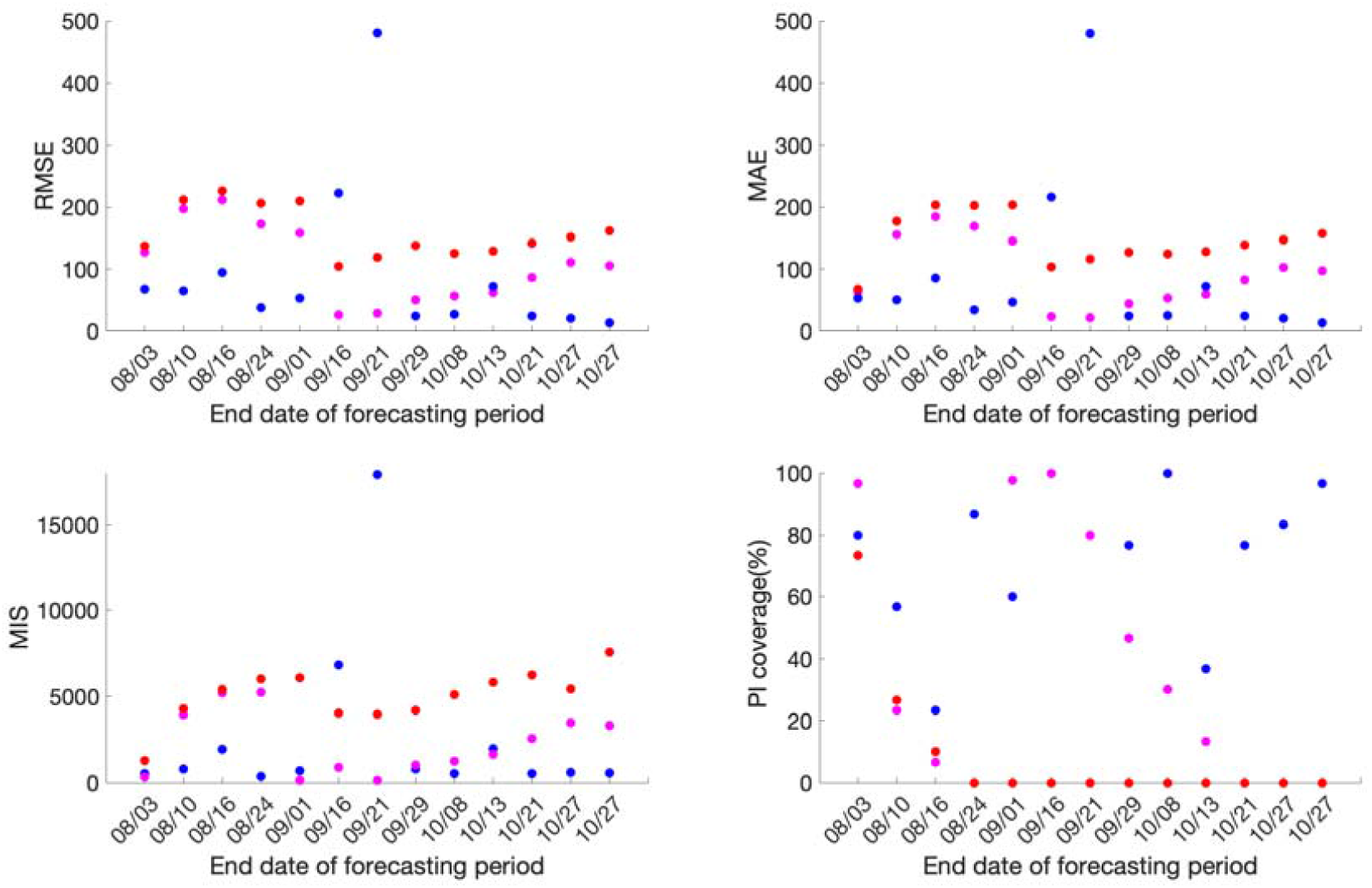
Forecasting period performance metrics for each of the thirteen sequential forecasting phases for GLM (magenta), Richards (red) and sub-epidemic (blue) model for Mexico. High 95% PI coverage and lower mean interval score (MIS), root mean square error (RMSE) and mean absolute error (MAE) indicate better performance.

Similarly, for Mexico City, the sub-epidemic model consistently outperforms the GLM and Richards growth model for ten of the thirteen forecasting phases in terms of RMSE and MAE and eleven forecasting phases in terms of the MIS. Whereas, in terms of 95% PI coverage, forecasting phases 08/31-09/29, 09/08-10/08 and 09/21-10/21 show zero 95% PI coverage across all three models. The sub-epidemic model outperforms the Richards and GLM model in six forecasting phases, with the Richards model performing better than the GLM model for the remaining four forecasting phases in terms of the 95% PI coverage (Fig 5, Table S3).

**Fig 5:**
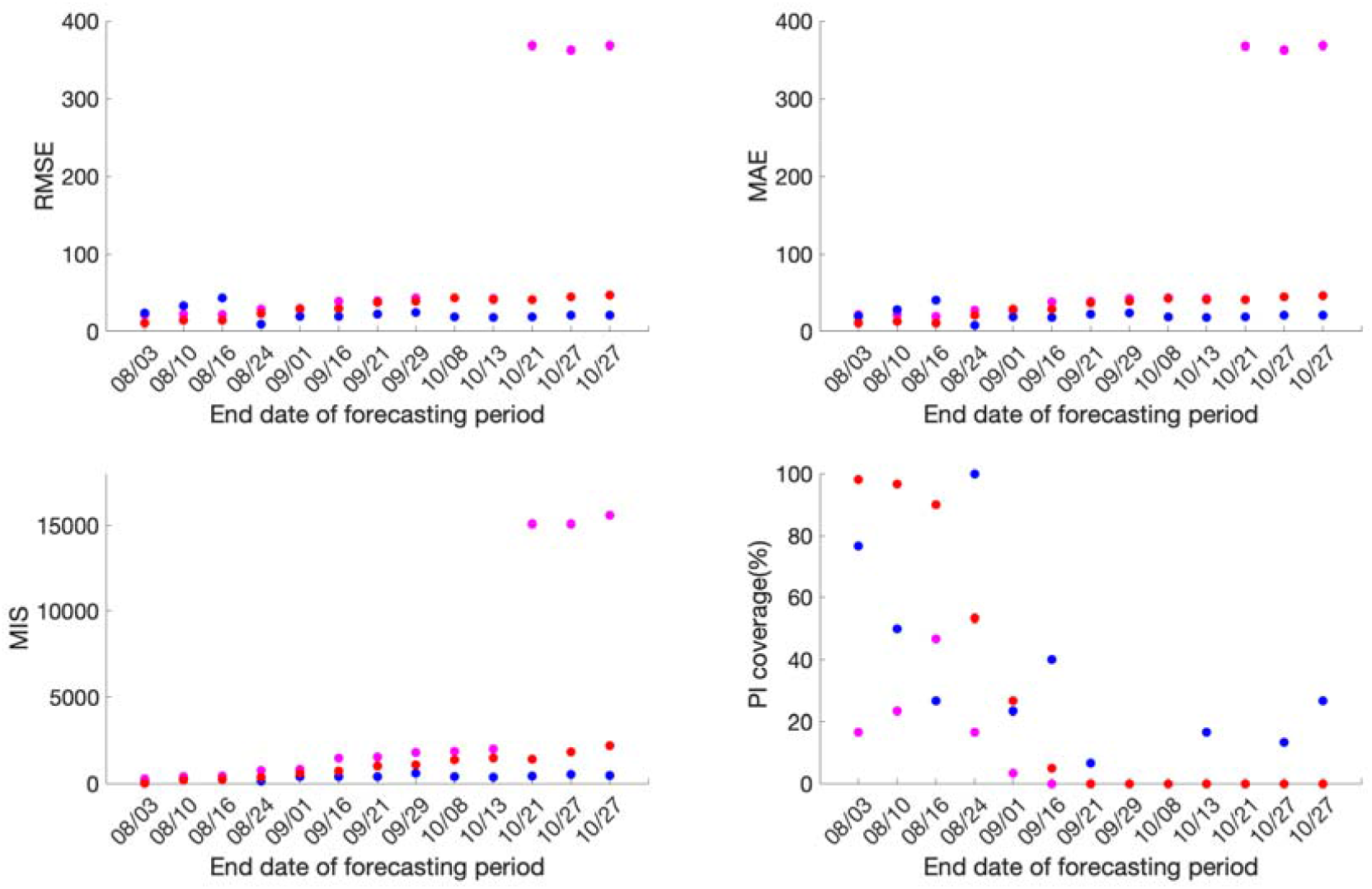
Forecasting period performance metrics for each of the thirteen sequential forecasting phases for GLM (magenta), Richards (red) and sub-epidemic (blue) model for the Mexico City. High 95% PI coverage and lower mean interval score (MIS), root mean square error (RMSE) and mean absolute error (MAE) indicate better performance.

### Comparison of daily death forecasts

The thirteen sequentially generated daily death forecasts from GLM and Richards growth model, for Mexico and Mexico City indicate towards a sustained decline in the number of deaths (S1 Fig, S2 Fig, S3 Fig and S4 Fig). However, the IHME model forecasts (retrieved from smoothed death data estimates, current projections scenario) indicate a decline in the number of deaths for the first six forecasts periods followed by a stable epidemic trajectory for the last seven forecasts, for Mexico City and Mexico. Unlike the GLM and Richards models, the sub-epidemic model is able to reproduce the observed stabilization of daily deaths observed after the first six forecast periods for Mexico and the last three forecast periods for Mexico City (S5 Fig, S6 Fig, S7 Fig and S8 Fig)

### Comparison of cumulative mortality forecasts

The total number of COVID-19 deaths is an important quantity to measure the progression of an epidemic. Here we present the results of the estimated cumulative death counts obtained from our 30-day ahead cumulative forecasts generated using the GLM, Richards and sub-epidemic growth model. We compare these results with the total mean smoothed death data estimates obtained from the three IHME modeling scenarios; current projection, universal masks and mandates easing. The total mean smoothed death data estimates obtained from the IHME current projections scenario as of November 11, 2020 are considered as a proxy for the actual death count for each date that the cumulative forecast is obtained (Figs 6 and 7).

**Fig 6:**
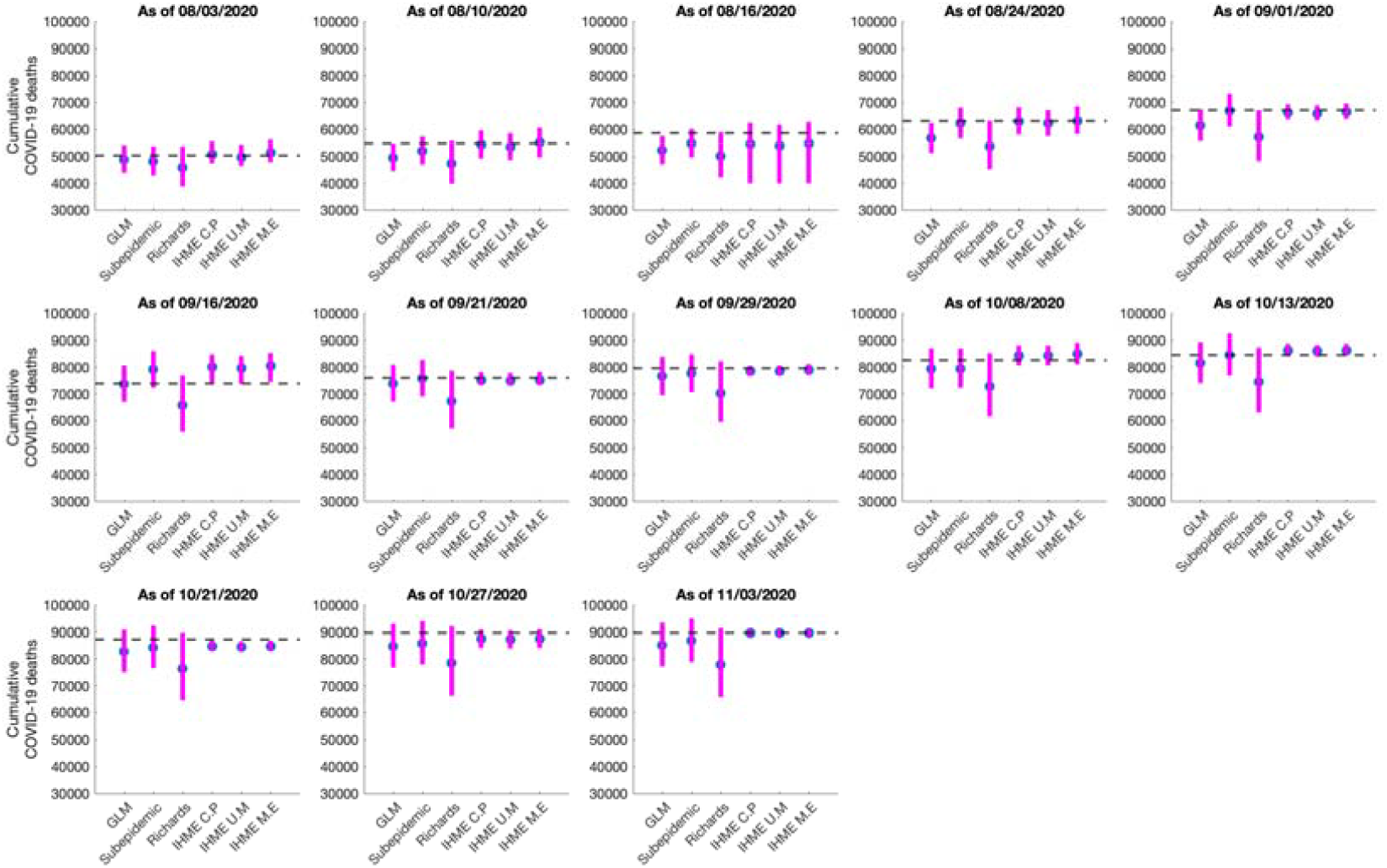
Systematic comparison of the six models (GLM, Richards, sub-epidemic model, IHME current projections (IHME C.P), IHME universal masks (IHME U.M) and IHME mandates easing (IHME M.E)) to predict the cumulative COVID-19 deaths for Mexico in the thirteen sequential forecasts. The blue circles represent the mean deaths and the magenta vertical line indicates the 95% PI around the mean death count. The horizontal dashed line represents the actual death count reported by that date in the November 11, 2020 IHME estimates files.

**Fig 7:**
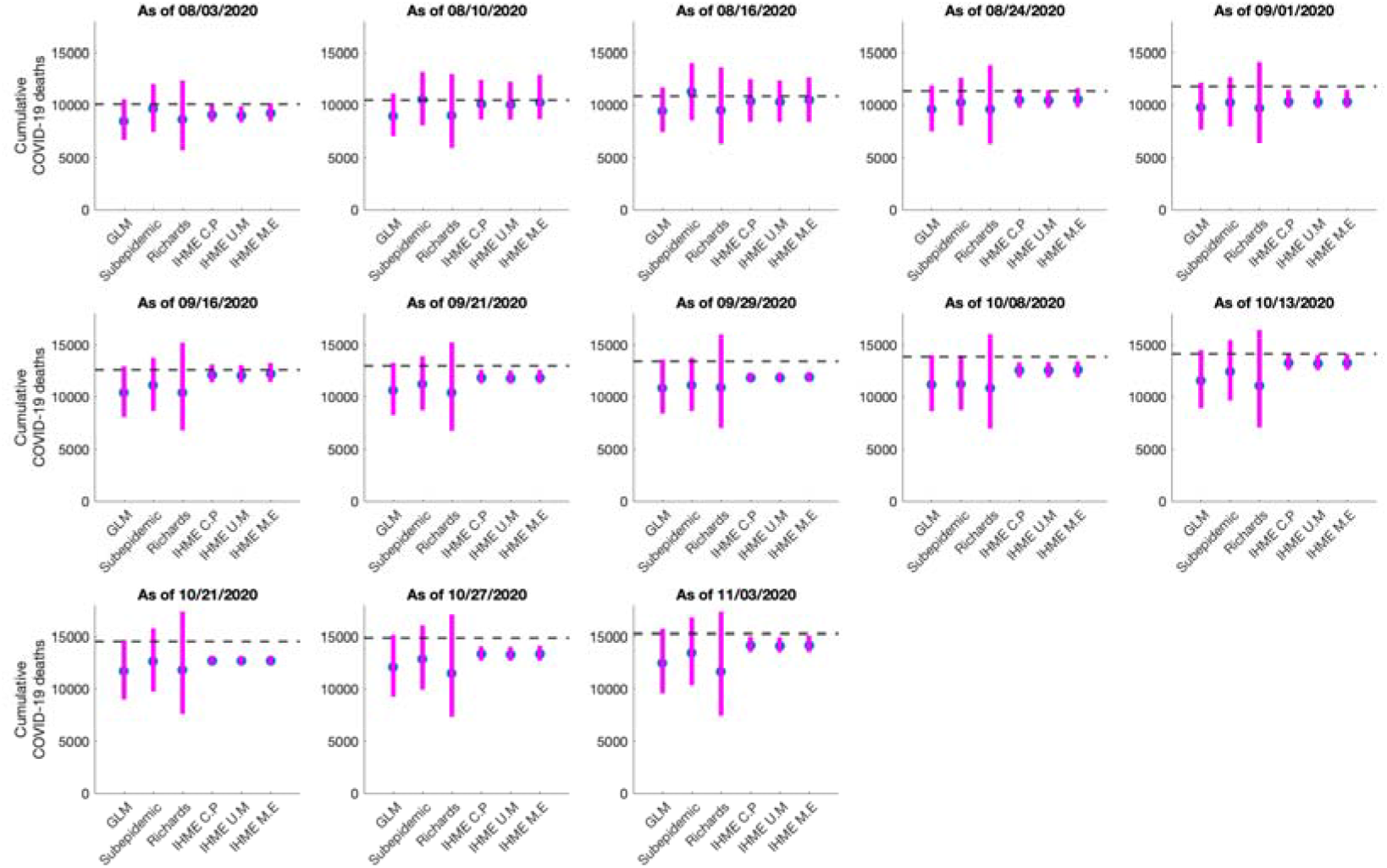
Systematic comparison of the six (GLM, Richards, sub-epidemic model, IHME current projections (IHME C.P), IHME universal masks (IHME U.M) and IHME mandates easing (IHME M.E))to predict the cumulative COVID-19 deaths for the Mexico City in the thirteen sequential forecasts. The blue circles represent the mean deaths and the magenta vertical line indicates the 95% PI around the mean death count. The horizontal dashed line represents the actual death count reported by that date in the November 11, 2020 IHME estimates files.

### Mexico

The 30-day ahead cumulative forecast results for the thirteen sequentially generated forecasts for Mexico utilizing GLM, Richards growth model, sub-epidemic growth model and the IHME model (current projections scenario) are presented in S9 Fig, S10 Fig, S11 Fig and S12 Fig. The cumulative mortality estimates comparison is given in Fig 6. For the first, second, third and thirteenth generated forecasts the GLM, sub-epidemic model and the Richards model tend to underestimate the true deaths counts (∼50,255, ∼54,857, ∼58,604, 89,730 deaths respectively), whereas the three IHME forecasting scenarios closely estimate the actual death counts for the first, second and thirteenth forecasting periods. For the fourth, fifth and seventh generated forecast the sub-epidemic model and the IHME scenarios most closely approximate the actual death counts (∼63,078, ∼67,075, ∼76,054 deaths respectively). For the sixth generated forecast the GLM model closely approximates the actual death count (∼73,911 deaths) whereas for the tenth generated forecast the sub-epidemic model closely approximates the actual deaths (∼84,471 deaths). For the eighth, ninth, eleventh and twelfth generated forecast GLM, Richards and sub-epidemic model tend to under-predict the actual death counts with the IHME model underestimating the actual death counts for eleventh and twelfth generated forecast and overestimating the total death counts for the ninth generated forecast (Table 2).

**Table 2:**
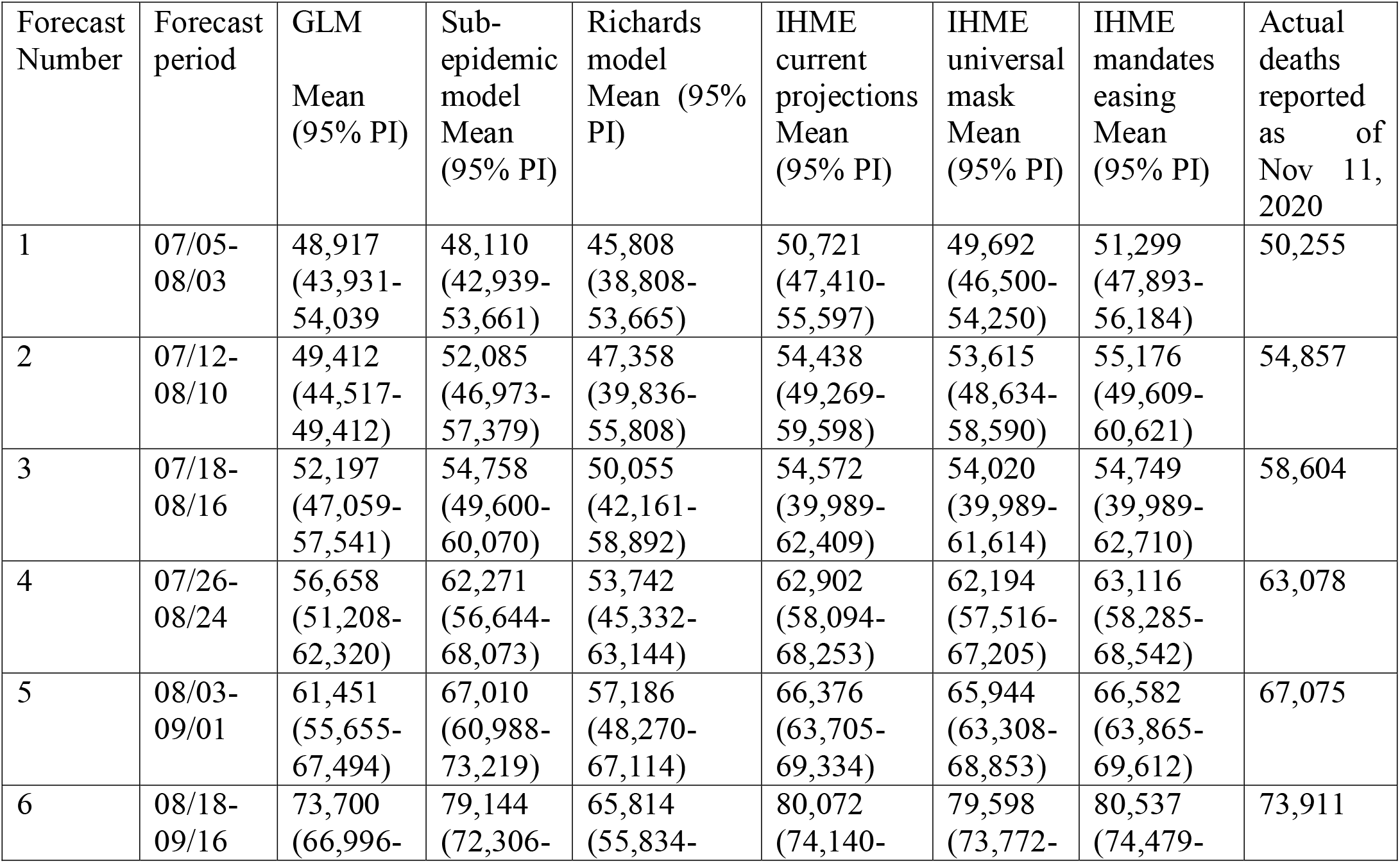
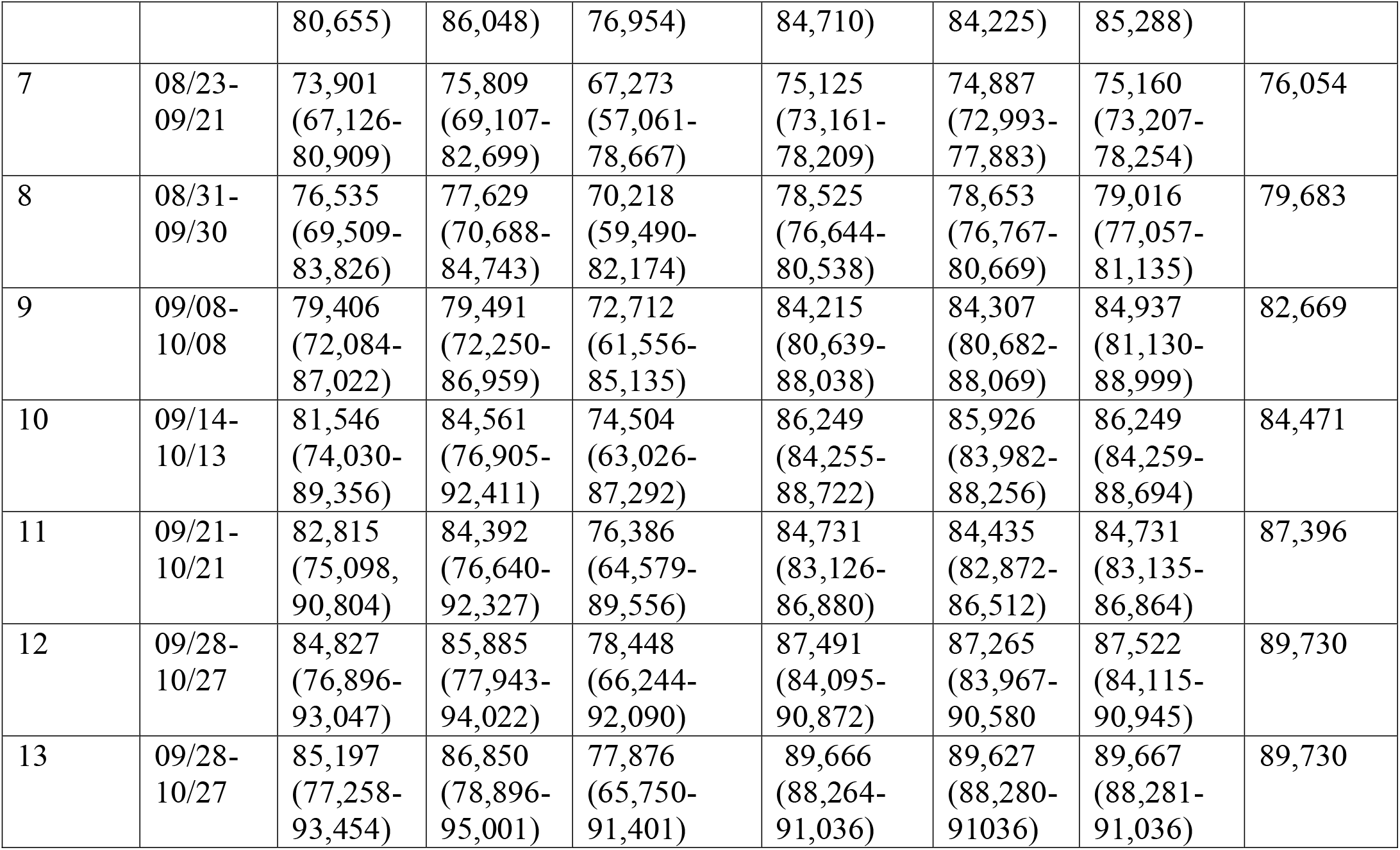
Cumulative mortality estimates obtained from the six models (GLM, Richards model, sub-epidemic model, IHME current projections, IHME universal mask and IHME mandates easing) for each forecasting period of the COVID-19 pandemic in Mexico (2020).

In summary, the Richards growth model consistently under-estimates the actual death count compared to the GLM, sub-epidemic model and three IHME model scenarios. The GLM model also provides lower estimates of the mean death counts compared to the sub-epidemic and three IHME model scenarios, but higher mean death estimates compared to the Richards model. The 95% PI for the Richards model is substantially wider than the other five models indicating wider uncertainty in the results. The actual mean death counts lie within the 95% PI of the sub-epidemic model for all the thirteen forecasts. Moreover, the three IHME model scenarios predict approximately similar cumulative death counts across the thirteen generated forecasts, indicating that the three scenarios do not differ substantially.

### Mexico City

The 30 day ahead cumulative forecast results for thirteen sequentially generated forecasts for the Mexico City utilizing GLM, Richards model, sub-epidemic growth model and IHME model (current projections scenario) are presented in S13 Fig, S14 Fig, S15 Fig and S16 Fig. The cumulative death comparison is given in Fig 7 and Table 3. For the first generated forecast the sub-epidemic model closely approximates the actual death count (∼10,081 deaths). For the second generated forecast, the sub-epidemic model and the IHME scenarios closely approximate the actual death count (∼10,496 deaths). For the third and sixth generated forecast GLM and Richards model underestimate the actual death count (∼10,859, ∼12,615 deaths respectively) whereas the sub-epidemic model closely estimates the death count for the third forecast and under-predicts the death count for the sixth forecast. The three IHME model scenarios seem to predict the actual death counts closely. For the fourth, fifth, seventh, eighth, ninth, tenth, eleventh, twelfth and thirteenth generated forecasts all models under-predict the actual death counts.

**Table 3:**
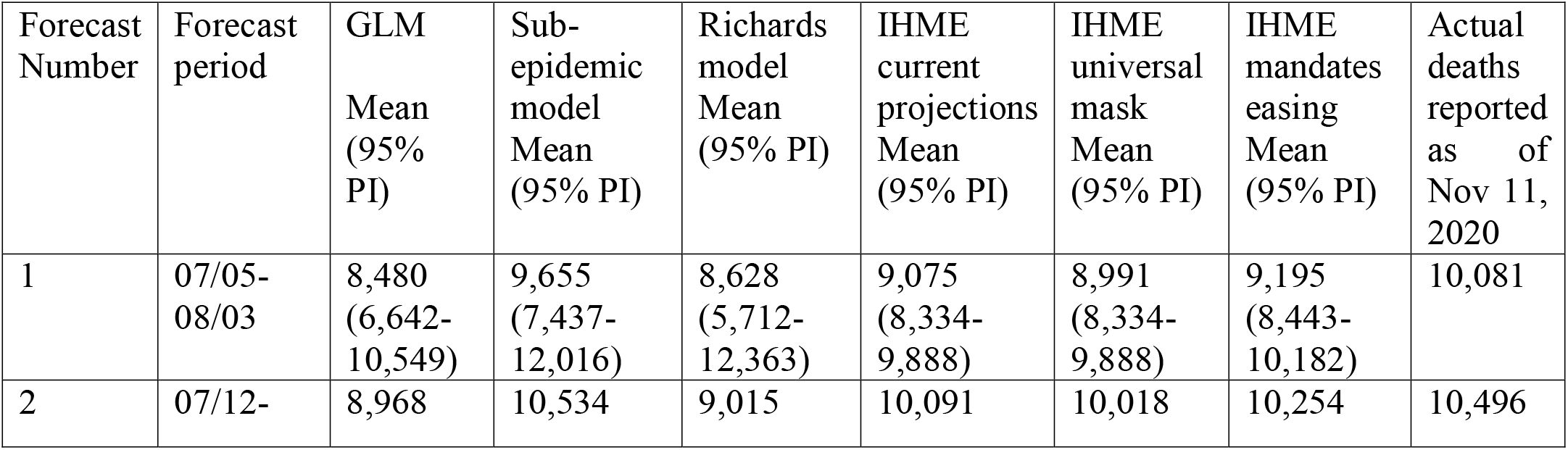
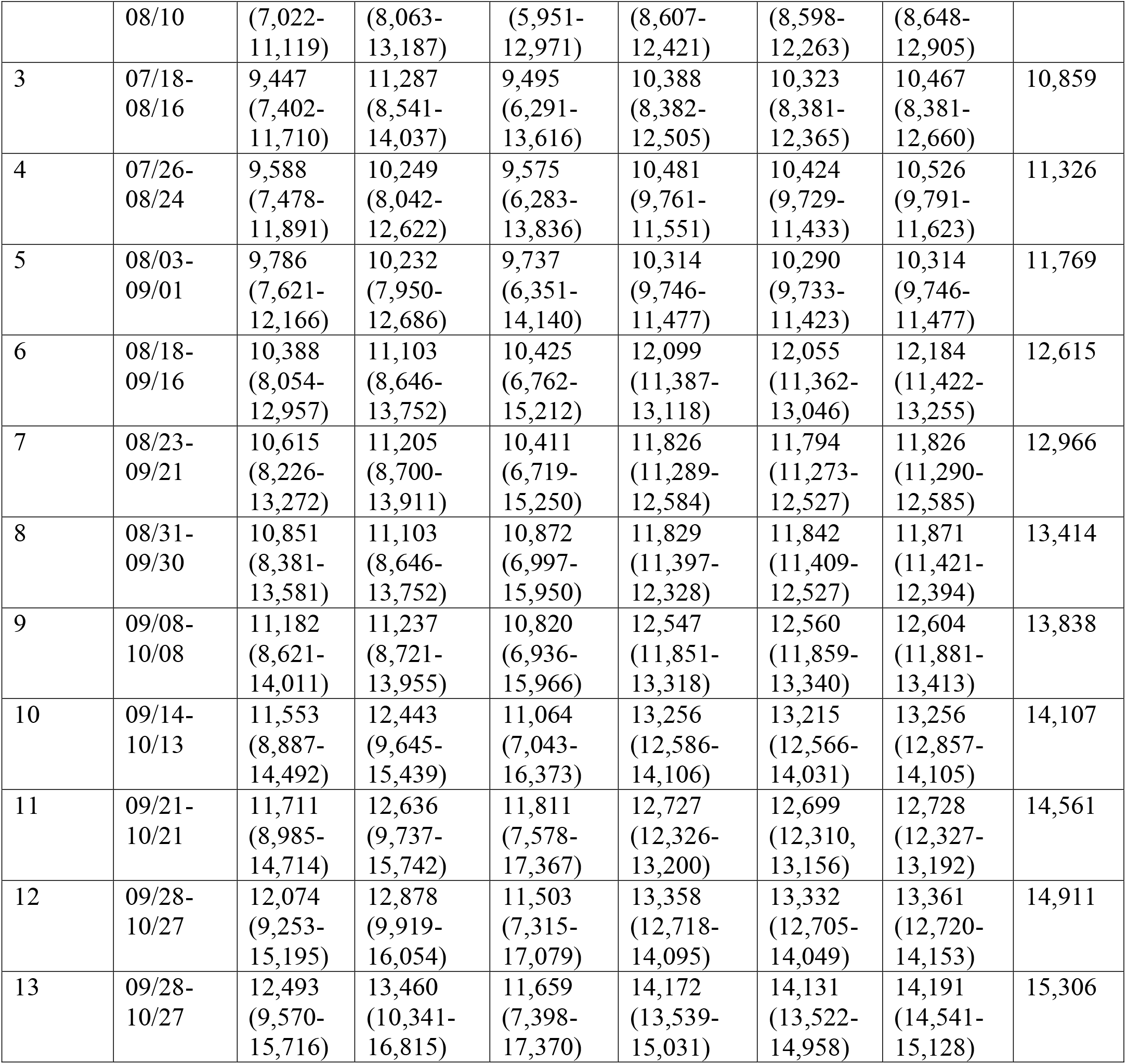
Cumulative mortality estimates obtained from the six models (GLM, Richards model, sub-epidemic model, IHME current projections, IHME universal mask and IHME mandates easing) for each forecasting period of the COVID-19 pandemic in Mexico City (2020).

In general, the Richards growth model has much wider 95% PI coverage compared to the other models indicating greater uncertainty in the results. The mean cumulative death count estimates for the GLM and Richards model closely approximate each other. However, the actual mean death counts lie within the 95% PI of the GLM and sub-epidemic model for all the thirteen forecasts. The three IHME model scenarios predict approximately similar cumulative death counts across the thirteen generated forecasts with much narrow 95% PI’s, indicating that three scenarios do not different substantially.

### Reproduction number

#### Estimate of reproduction number, R from genomic data analysis

The majority of analyzed Mexican SARS-CoV-2 sequences (69 out of 83) have been sampled in March and April 2020. These sequences are spread along the whole global SARS-CoV-2 phylogeny (Fig 8) and split into multiple clusters. This indicates multiple introductions of SARS-CoV-2 to the country during the initial pandemic stage (February 27-May 29, 2020). For the largest cluster of size 42, the reproduction number was estimated at *R* = 1.3 (95% HPD interval [1.1,1.5]).

**Fig 8:**
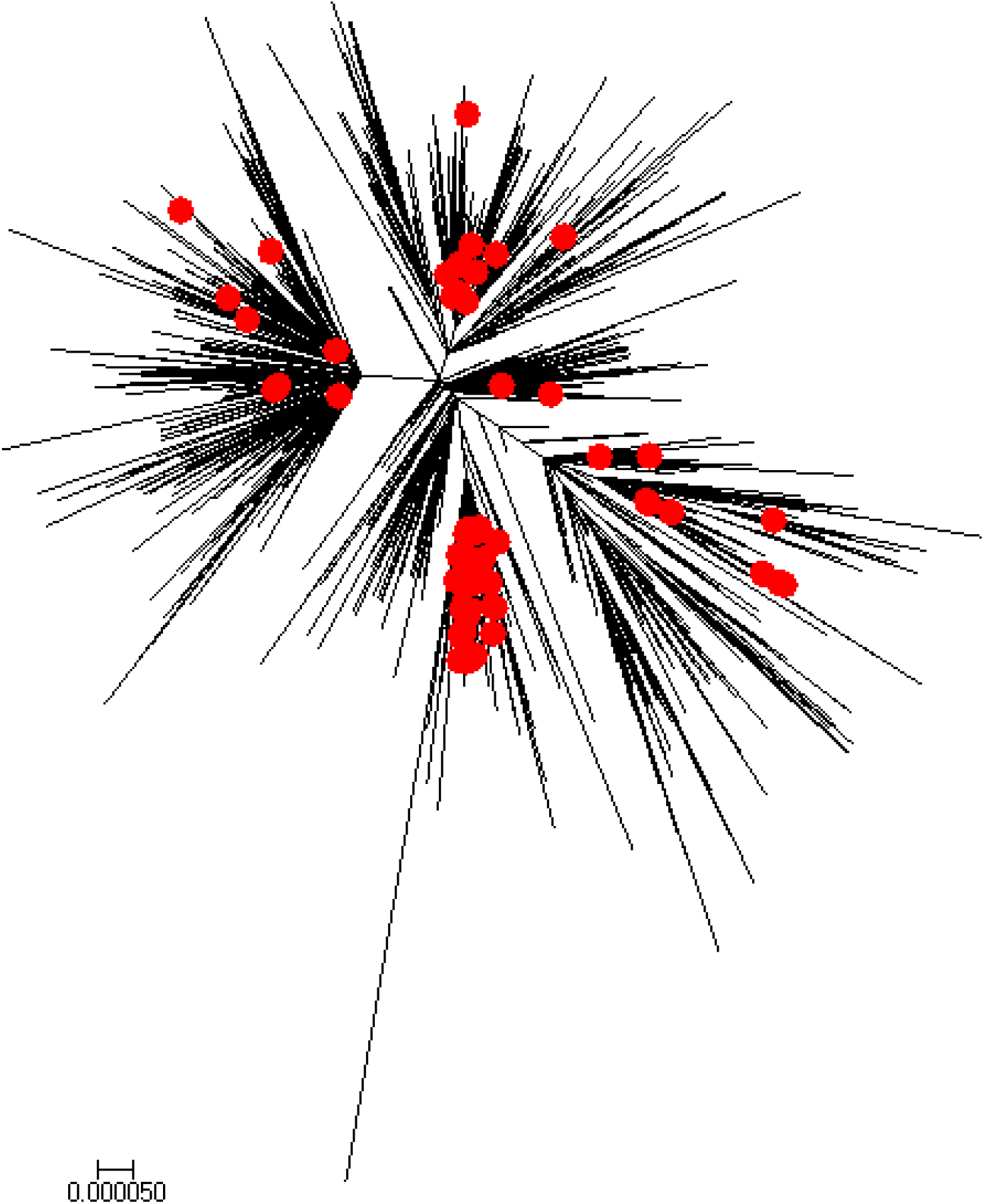
Global neighbor-joining tree for SARS-CoV-2 genomic data from February 27-May 29, 2020. Sequences sampled in Mexico are highlighted in red.

#### Estimate of reproduction number, *R*_*t*_ from case incidence data

The reproduction number from the case incidence data (February 27-May 29, 2020) using GGM was estimated at *R*_*t*_∼1.1(95% CI:1.1,1.1), in accordance with the estimate of *R*_*t*_ obtained from the genomic data analysis. The growth rate parameter, *r*, was estimated at 1.2 (95% CI: 1.1, 1.4) and the deceleration of growth parameter, *p*, was estimated at 0.7 (95% CI: 0.68,0.71), indicating early sub-exponential growth dynamics of the epidemic (Fig 9).

**Fig 9:**
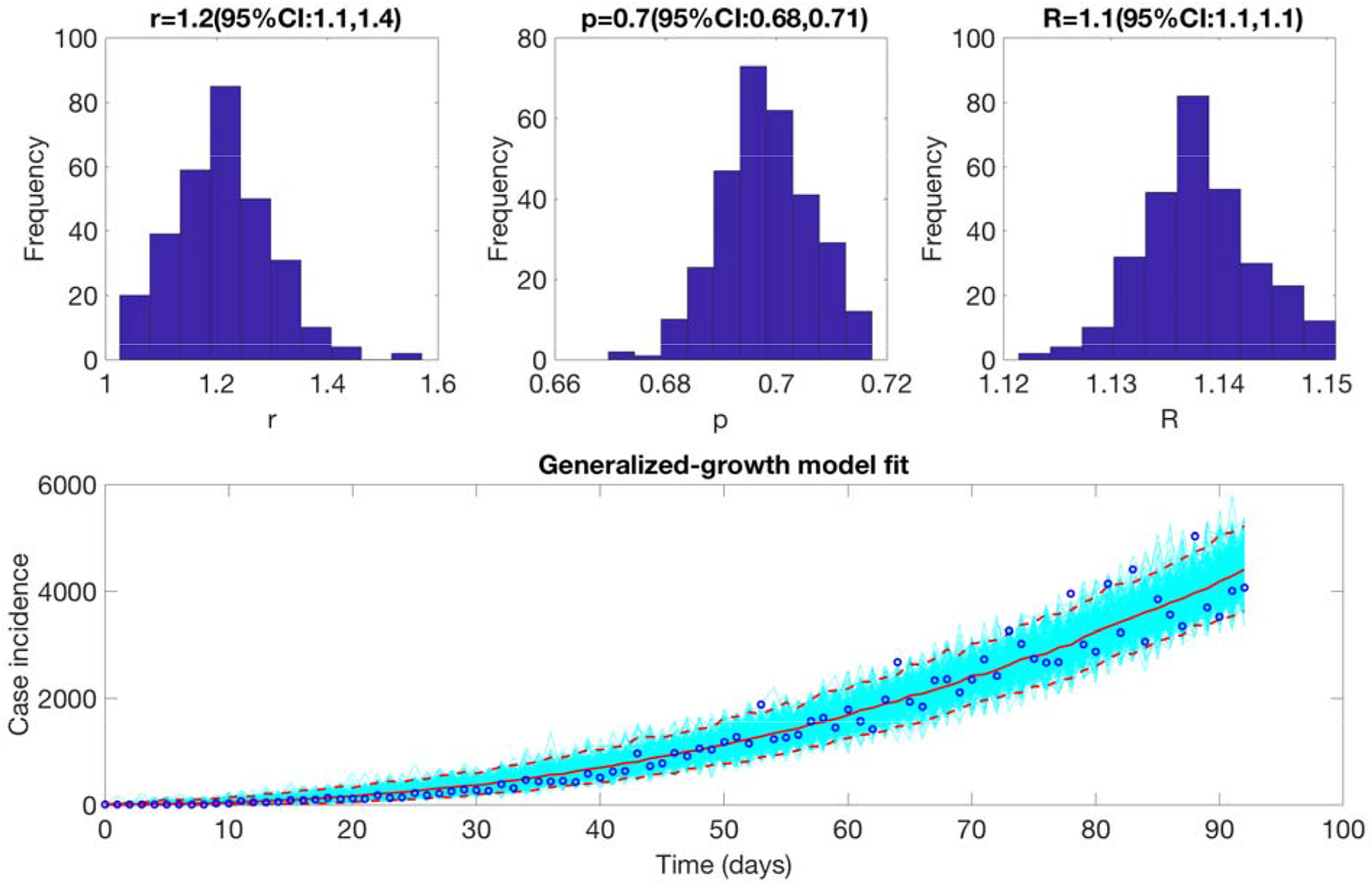
Upper panel: Reproduction number with 95% CI estimated using the GGM model. The estimated reproduction number of the COVID-19 epidemic in Mexico as of May 29, 2020, 2020 is 1.1 (95% CI: 1.1, 1.1). The growth rate parameter, r, is estimated at 1.2(95%CI: 1.1, 1.4) and the deceleration of growth parameter, p, is estimated at 0.7 (95%CI:0.68, 0.71). Lower panel: The lower panel shows the GGM fit to the case incidence data for the first 90 days.

#### Estimate of instantaneous reproduction number, *R*_*t*_

The instantaneous reproduction number for Mexico remained consistently above 1.0 until the end of May 2020, after which the reproduction number has fluctuated around 1.0 with the estimate of *R*_*t*_∼0.93 (95% CrI: 0.91, 0.94) as of September 27, 2020. For Mexico City, the reproduction number remained above 1.0 until the end of June after which it has fluctuated around 1.0 with the estimate of *R*_*t*_*∼*0.96 (95% CrI: 0.93, 0.99) as of September 27, 2020 (Fig 10).

**Fig 10:**
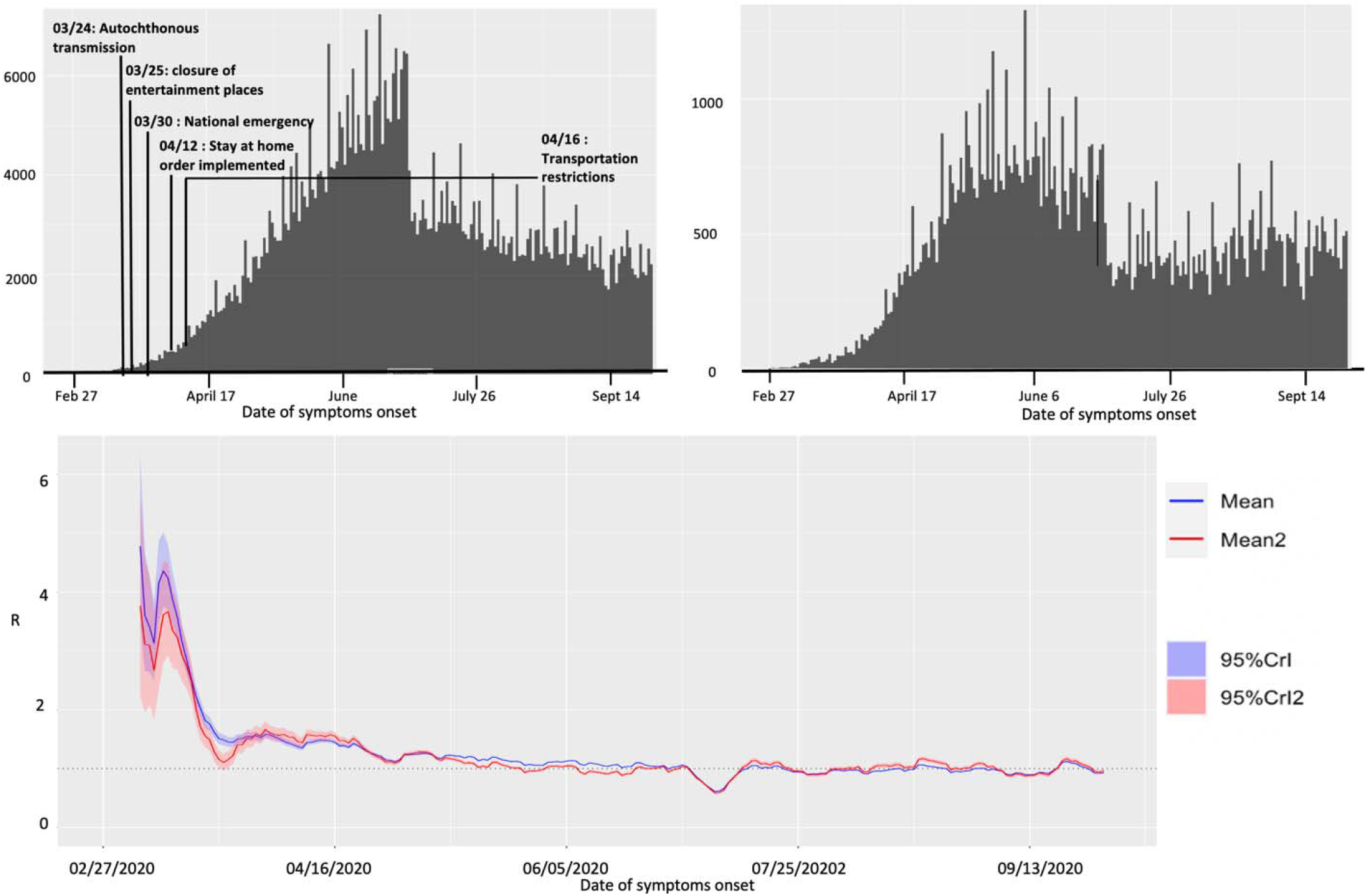
Upper panel: Epidemiological curve (by the dates of symptom onset) for Mexico (left panel) and Mexico City (right panel) as of September 27, 2020. Lower panel: Instantaneous reproduction number with 95% credible intervals for the COVID-19 epidemic in Mexico as of September 27, 2020. The red solid line represents the mean reproduction number for Mexico and the red shaded area represents the 95% credible interval around it. The blue solid line represents the mean reproduction number for the Mexico City and the blue shaded region represents the 95% credible interval around it.

### Spatial analysis

Fig S17 shows the result from pre-processing COVID-19 data into growth rate functions. The results of clustering are shown in Fig S18 as a dendrogram plot and the states color coded based on their cluster membership within the map of Mexico (Fig 11; left panel). The four predominant clusters we identified include the following states:

**Fig 11:**
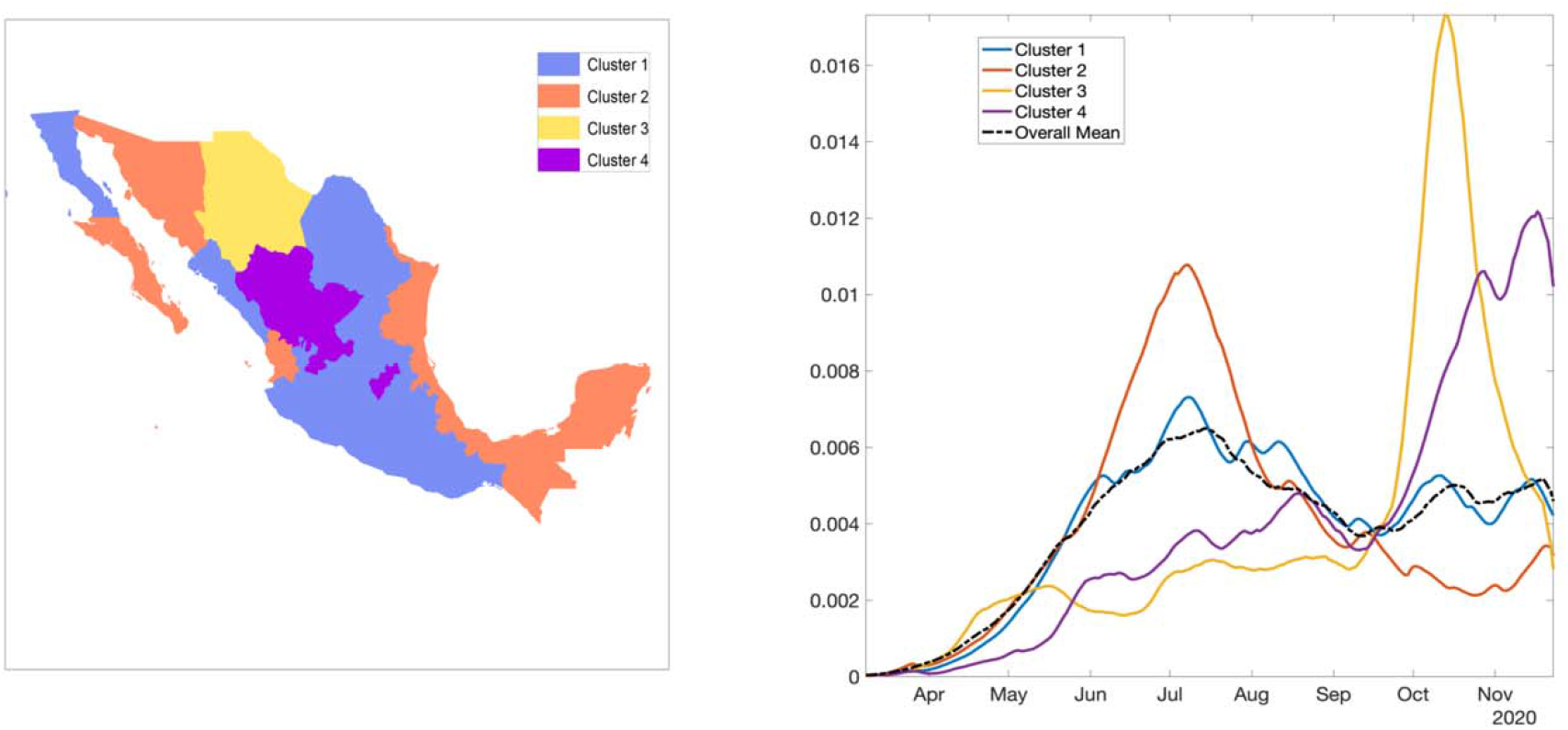
Clusters of states by their growth rates. Cluster 1 in blue, cluster 2 in orange, cluster 3 in yellow, and cluster 4 in purple. The right panel shows the average growth rate curves for each cluster (solid curves) and their overall average (black broken curve).

Cluster 1: Baja California, Coahuila, Colima, Mexico City, Guanajuato, Guerrero, Hidalgo, Jalisco, Mexico, Michoacán, Morelos, Nuevo Leon, Oaxaca, Puebla, San Luis Potosi, Sinaloa, and Tlaxcala

Cluster 2: Baja California Sur, Campeche, Chiapas, Nayarit, Quintana Roo, Sonora, Tabasco, Tamaulipas, Veracruz, and Yucatan

Cluster 3: Chihuahua

Cluster 4: Aguascalientes, Durango, Queretaro, and Zacatecas

Fig 11(right panel) shows the average shape of growth rate curves in each cluster and the overall average. Fig S19 shows mean growth rate curves and one standard-deviation bands around it, in each cluster. Since cluster 3 included only one state, average growth rate of cluster 1, cluster 2, and cluster 4 are shown. The average growth patterns in the three categories are very distinct and clearly visible. For cluster 1, the rate rises rapidly from April to July and then shows small fluctuations. For cluster 2, there is rapid increase in growth rate from April to July followed by a rapid decline. Chihuahua in cluster 3 shows a slow growth rate until September followed by a rapid rise until mid-September which then declines rapidly. For cluster 4, the rate rises slowly, from April to September, and then shows a rapid rise (Fig S20).

From the colormap (Fig 12) we can see that the cases were concentrated from the beginning in the central region in Mexico and Mexico City. Daily cases have been square root transformed to reduce variability in the amplitude of the time series while dashed lines separate the Northern, Central, and Southern regions. Fig S20 shows the timeseries graph of daily COVID-19 new cases by the date for all states, Northern states, Central states, and the Southern states. As observed for both Northern and Central regions including the national level, the epidemic peaked in mid-July followed by a decline at around mid-September, which then started rising again. Southern states exhibit a stable decline. Fig S21 shows the total number of COVID-19 cases at state level as of December 5, 2020. Some of the areas with a higher concentration of COVID-19 cases are: Mexico City, Mexico state, Guanajuato in the central region and, Nuevo Leon in the Northern region.

**Fig 12:**
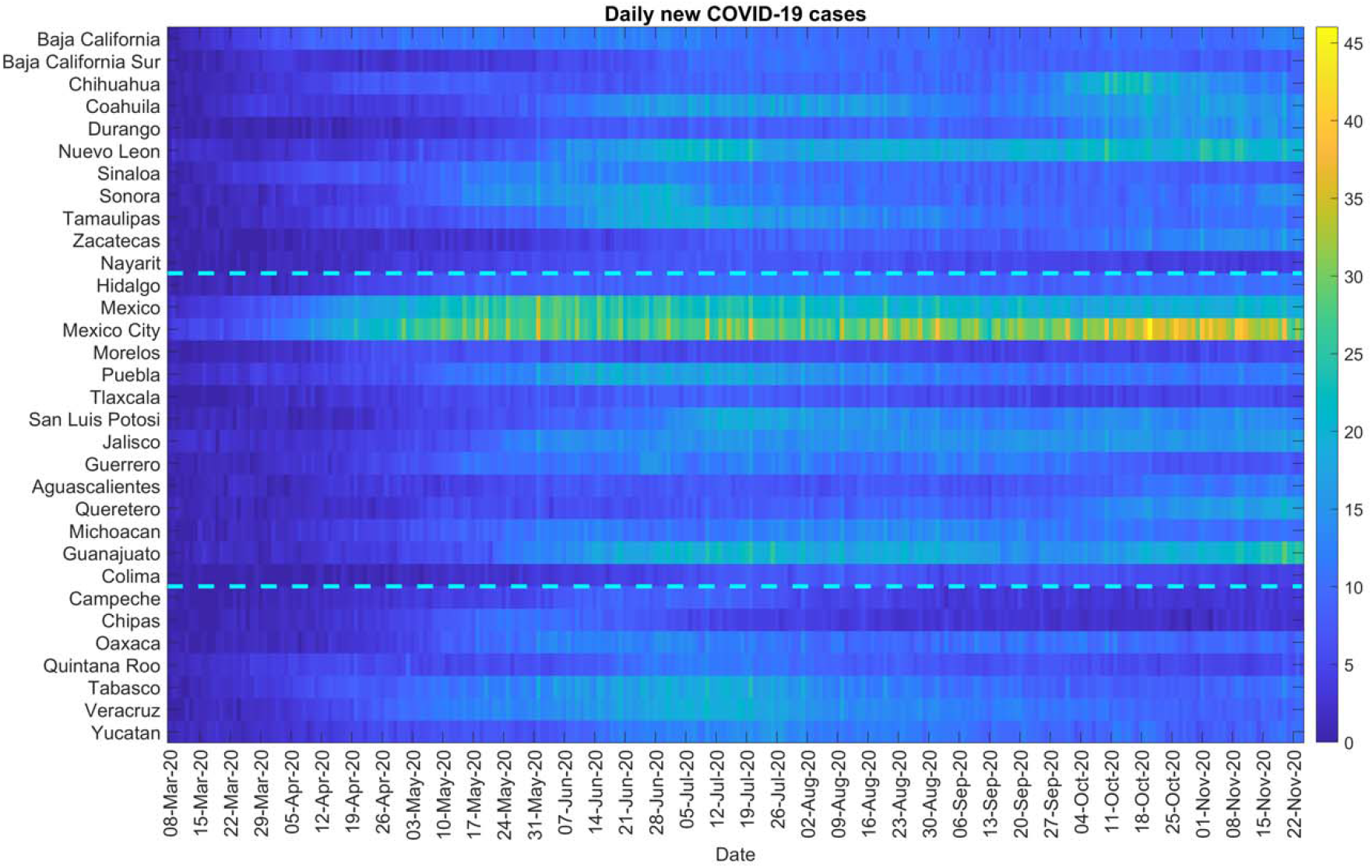
Color scale image of daily COVID-19 cases by region.

### Twitter data analysis

The epicurve for Mexico is overlaid with the curve of tweets indicating stay at home orders in Mexico as shown in S22 Fig. The engagement of people in Mexico with the #quedateencasa hashtag (stay-at-home order hashtag) has been gradually declining as the number of cases have continued to increase or remain at a steady pace, showing the frustration and apathy of public on lock downs and restrictions. Mostly the non-government public health experts are calling for more lockdowns or continued social distancing measures (without being heard by the authorities). It could also imply that the population is not following the government’s stay at home orders and hence we continue to observe the cases. S22 Fig shows that the highest number of tweets were made in the earlier part of the epidemic, with the number of tweets declining as of mid-May 2020. In contrast, the number of cases by onset dates peaked around mid-June. The correlation coefficient between the epi-curve of cases by dates of onset and the curve of tweets representing the stay-at-home orders was estimated at R=-0.001 from March 12-November 11, 2020.

## Discussion

The results of our GLM model fit to the smoothed death data estimates for all the thirteen calibration phases and GGM fit to the case incidence data indicate sub-exponential growth dynamics of COVID-19 epidemic in the Mexico and Mexico City with the parameter *p* estimated between (*p* ∼0.6-0.8). Moreover, the early estimates of *R* indicate towards sustained disease transmission in the country. As *R*_*t*_ fluctuates around ∼1.0 since the end of July 2020, we observe different epidemic growth patterns at the national and state level. With ongoing virus transmission in Mexico, the Twitter analysis implies the relaxation of lockdowns, with inconsequential decline in the mobility patterns observed over the last few weeks as evident from the Apple’s mobility trends. Moreover, the systematic comparison of our models across thirteen sequential forecasts deems sub-epidemic model as the most appropriate model for mortality forecasting. The sub-epidemic model is also able to reproduce the stabilization in the trajectory of mortality forecasts as observed by the IHME forecasts.

The sub-exponential growth pattern of the COVID-19 epidemic in Mexico can be attributed to a myriad of factors including non-homogenous mixing, spatial structure, population mobility, behavior changes and interventions [87]. Our results are consistent with the sub-exponential growth patterns of COVID-19 outbreaks observed in Mexico [88] and Chile [89]. Along with the observed sub-exponential growth dynamics of the COVID-19 epidemic in Mexico, the reproduction number estimated from the genomic sequence analysis and the case incidence data (*R*_*t*_∼ 1.1-1.2) indicate towards sustained transmission of SARS-CoV-2 in Mexico during the early transmission phase of the virus (February 27-May 29, 2020). Our estimates of *R*_*t*_ are similar to the estimates of reproduction number retrieved from other studies conducted in Mexico [90], Chile [91, 92], Peru [93] and Brazil [94]. The early estimate of *R*_*t*_ obtained from the Cori et al. method in our study also coincides with the early estimates of *R*_*t*_ obtained from the case incidence data and the genomic data (*R*_*t*_∼1). The instantaneous reproduction number estimated from our study shows that *R*_*t*_ is slightly above 1 since the end of March, without a significant increase. This is in accordance with the estimates of *R*_*t*_ obtained from a study conducted in Mexico [95].

In general, Mexico has seen a sustained SARS-CoV-2 transmission and an increase or a sustained number of cases despite the implementation of the social distancing interventions including the stay-at-home orders that were eased around June 2020. As our twitter data analysis also shows, the number of cases by onset dates was negatively correlated to the stay-at-home orders. A possible explanation indicates that people might have stopped following the government’s preventive orders to stay at home as a result of pandemic fatigue [96, 97]. Mexico has been one of the countries where the stay-at-home orders have been least respected. The average reduction in mobility in Mexico is reported to be ∼35.4% compared to 71% mobility reduction in Brazil, and 86% mobility reduction in Argentina and Colombia [98]. These preventive orders have affected the Mexican population disproportionately, with some proportion of the population exhibiting aggression towards quarantine and stay-at-home orders [36]. However, the public health professionals are frustrated towards the relaxation of stay-at-home orders and reopening of the country, as the cases and deaths keep mounting. We can also appreciate the variable spatial-temporal dynamics of the COVID-19 epidemic in Mexico. Our classification of epidemic pattern at the state level in Mexico shows distinct variation of growth rates across states. For instance, cluster 1 including Baja California, Colima and Mexico City has stable growth at a higher rate and cluster 4 including Aguascalientes, Durango, Queretaro, and Zacatecas shows a rising pattern in the growth rate (Figure 11). This information can be utilized by the states in guiding their decision regarding the implementation of public health measures. For example, states in cluster 1 and 4 may need strict public health measures to contain the epidemic.

Appropriate short-term forecasts can also help gauge the impact of interventions in near real-time. In this study we compared the performance of our three models for short-term real-time forecasting the COVID-19 mortality estimates in Mexico and Mexico City. As in Figs 2-5 the sub-epidemic model can be declared the most appropriate model as it exhibits the most desirable performance metrics across majority of the calibration and forecasting phases. This model has the capacity to accommodate more complex epidemic trajectories suggesting a longer epidemic wave and can better adjust to the early signs of changes in disease transmission, while other models (GLM and Richards) are less reactive. This model can also be utilized as a potential forecasting tool for other cities in Mexico; comparing its results with other prediction models. Moreover, further short-term forecasts (5,10 days) could be also be conducted with the sub-epidemic model using the consecutive calibration phases to reduce the error metrics [52].

Overall, the sequential forecasts based on the daily smoothed death estimates for Mexico from the two models (GLM, Richards) suggest a decline in over-all deaths (S1 Fig and S2 Fig) consistent with the sustained decline in COVID-19 associated case fatalities since mid-August as reported by the Mexican government, officially [99]. However, this decline in COVID-19 deaths can be attributed to the inaccurate reporting of deaths in the surveillance system or downplay of fatalities by the government. For instance, the reported excess deaths as of September 26, 2020 are estimated to be 193170, with 139151 deaths attributable to COVID-19 [100]. While the official tally of COVID-19 deaths in Mexico is only exceeded by USA and Brazil, its roughly approximated with that of India, a country whose population is ten times larger than Mexico [101]. As observed earlier, the easing of the social distancing interventions and lifting of lockdowns in Mexico in the month of June led to a surge of the COVID-19 associated deaths [102]. In June, the government of Mexico also inaccurately forecasted that a potential decline in the number of COVID-19 deaths will be observed by September, 2020 [103]. Therefore, the forecasting trends need to be interpreted cautiously in order to inform policies. The IHME model also shows a decline in COVID-19 deaths in Mexico from mid-August-September, which have stabilized since then for the last six forecast periods (S5 Fig). The sub-epidemic model also indicates a stabilization of the deaths for the last seven forecast periods (S6 Fig) consistent with the results obtained from the IHME model.

Similarly, for Mexico City, the sequential forecasts obtained from the GLM and Richards model fit to the daily death data estimates indicate a decline in the overall deaths (S3 Fig, S4 Fig). The IHME and sub-epidemic models on the other hand indicate a stabilized trajectory of deaths for the last three forecast periods (S7 Fig and S8 Fig) (suggesting that the actual death counts might not be decreasing in Mexico City) as seen with Mexico. Based on the death data, the observed decline or stability in death predictions could likely reflect the false slowing down of the epidemic in Mexico City [102]. Moreover, insufficient testing can also result in an inaccurate trajectory of the COVID-19 mortality curve [104].

The cumulative comparison of deaths in Mexico and Mexico City indicates that in general, the Richards model has under-performed in predicting the actual death counts with much wider uncertainty around the mean death estimates. The Richards model has also failed to capture the early sub-exponential dynamics of the mortality curve. The cumulative death counts obtained from the flexible sub-epidemic model closely approximate the total mean death counts obtained from the three IHME modeling scenarios. Whereas the GLM slightly under predicts the cumulative death counts (Fig 6, Fig 7). On the other hand, another competing model, the COVID-19 predictions model projects 87,151 deaths (95% PI:84,414, 91,883) for Mexico as of October 27, 2020 (last forecasting phase), an estimate that closely approximates the estimates obtained from the GLM model (between 77,258-93,454 deaths) [105].

The three models (GLM, Richards, sub-epidemic model) used in this study generally provide good fits to the mortality curve based on the residuals. The Richards model however is unable to capture the early sub-exponential dynamics of the mortality curve. Moreover, these phenomenological models are particularly valuable for providing rapid predictions of the epidemics in complex scenarios that can be used for real-time preparedness since these models do not require specific disease transmission processes to account for the interventions. Since these models do not explicitly account for behavioral changes, the results should be interpreted with caution. Importantly, since the mortality curves employed in this study are reported according to the date of reporting, they are likely influenced by variation in the testing rates and related factors including the case fatality rates. Further, delays in reporting of deaths due to the magnitude of the epidemic could also influence our predictions. Moreover, using the reporting date is not ideal due to the time difference between the date of death and the reporting date of death, which at a given moment can give a false impression of the ongoing circumstances.

This paper is not exempt from limitations. First, the IHME (current projections, mandated mask, and worst-case scenario) model utilized has been revised multiple times over the course of the pandemic and differs substantially in methodology, assumptions, range of predictions and quantities estimated. Second, the IHME has been irregular in publishing the downloadable estimates online for some periods. Third, we model the death estimates by date of reporting rather than by the date of death. Lastly, the unpredictable social component of the epidemic on ground is also a limiting factor for the study as we do not know the ground truth mortality pattern when the forecasts are generated.

In conclusion, the reproduction number has been fluctuating around ∼1.0 since the end of July-end of September 2020, indicating sustained virus transmission in the region. Moreover, the country has seen much lower mobility reduction and mixed reactions towards the stay-at-home orders contributing towards the virus transmission in the country. Moreover, the spatial analysis indicates that states like Mexico, Michoacán, Morelos, Nuevo Leon, Baja California need to put in place stronger public health strategies to contain the rising patterns in growth rates. The GLM and sub-epidemic model applied to mortality data in Mexico provide reasonable estimates for short-term projections in near real-time. While the GLM and Richards models predict that the COVID-19 outbreak in Mexico and Mexico City may be on a sustained decline, the sub-epidemic model and IHME model predict a stabilization of daily deaths. However, our forecasts need to be interpreted with caution given the dynamic implementation and lifting of the social distancing measures.

## Data Availability

Data for short term forecasts is available from the IHME downloadable estimates
Data for mobility trends is available from the Apple mobility trends
Genomic data is available from the GSAID repository
Case incidence data is available from the Ministry of Health, Mexico

http://www.healthdata.org/news-release/new-ihme-covid-19-forecasts-show-deaths-mexico-exceeding-50000

https://covid19.apple.com/mobility

https://www.gisaid.org/

https://www.gob.mx/salud/documentos/datos-abiertos-152127

## Author Contributions

Conceptualization, G.C. and A.T.; methodology, G.C, A.T.; validation, G.C.; formal analysis, A.T., G.C.; investigation, A.T. S.D., J.M.B., P.S.; resources, G.C., A.T. J.M.B., B.E. A.B, P.S; data curation, A.T.; writing—original draft preparation, A.T., G.C.; writing, review and editing, A.T., G.C., J.M.B., P.S., S.D., C.C.G, B.E., N.G.B., R.A.S, A.K, R.L, A.S, H.G., N.G.C., A.I.B., M.E.J; visualization, A.T., G.C.; supervision, G.C.; project administration, G.C.; funding acquisition, G.C. All authors have read and agreed to the published version of the manuscript.

## Supporting Information

S1 Fig: COVID-19 deaths forecasts using daily deaths, GLM model, Mexico: 30-days ahead forecasts based on the Generalized Logistic Growth Model (GLM) calibrated using an increasing amount of daily death data (blue circles): 107, 114, 120, 128, 136, 151, 156, 164, 172, 179, 185, 193, 193 epidemic days. The vertical dashed line indicates the end of the calibration period and start of the forecasting period. The mean (solid red line) and 95% PIs (dashed red lines) of the model fit and forecast are shown.

S2 Fig: COVID-19 death forecasts using daily deaths, Richards model, Mexico: 30-days ahead forecasts based on the Richards model calibrated using an increasing amount of daily death data (blue circles): 107, 114, 120, 128, 136, 151, 156, 164, 172, 179, 185, 193, 193 epidemic days. The vertical dashed line indicates the end of the calibration period and start of the forecasting period. The mean (solid red line) and 95% PIs (dashed red lines) of the model fit and forecast are shown

S3 Fig: COVID-19 death forecasts using daily deaths, GLM model, Mexico City: 30-days ahead forecasts based on the GLM model calibrated using an increasing amount of daily death data (blue circles): 107, 114, 120, 128, 136, 151, 156, 164, 172, 179, 185, 193, 193 epidemic days. The vertical dashed line indicates the end of the calibration period and start of the forecasting period. The mean (solid red line) and 95% PIs (dashed red lines) of the model fit and forecast are shown.

S4 Fig: COVID-19 death forecasts using daily deaths, Richards model, Mexico City: 30-days ahead forecasts based on the Richards model calibrated using an increasing amount of daily death data (blue circles): 107, 114, 120, 128, 136, 151, 156, 164, 172, 179, 185, 193, 193 epidemic days. The vertical dashed line indicates the end of the calibration period and start of the forecasting period. The mean (solid red line) and 95% PIs (dashed red lines) of the model fit and forecast are shown.

S5 Fig: COVID-19 death forecasts using daily deaths, IHME model, Mexico: 30-days ahead forecasts based on the IHME model calibrated using an increasing amount of daily death data (blue circles): 107, 114, 120, 128, 136, 151, 156, 164, 172, 179, 185, 193, 193 epidemic days. The vertical dashed line indicates the end of the calibration period and start of the forecasting period. The mean (solid red line) and 95% PIs (dashed red lines) of the model fit and forecast are shown.

S6 Fig: COVID-19 death forecasts using daily deaths, sub-epidemic wave model, Mexico: 30-days ahead forecasts based on the sub-epidemic wave model calibrated using an increasing amount of daily death data (blue circles): 107, 114, 120, 128, 136, 151, 156, 164, 172, 179, 185, 193, 193 epidemic days. The vertical dashed line indicates the end of the calibration period and start of the forecasting period. The mean (solid red line) and 95% PIs (dashed red lines) of the model fit and forecast are shown.

S7 Fig: COVID-19 death forecasts using daily deaths, IHME model, Mexico City: 30-days ahead forecasts based on the IHME model calibrated using an increasing amount of daily death data (blue circles): 107, 114, 120, 128, 136, 151, 156, 164, 172, 179, 185, 193, 193 epidemic days. The vertical dashed line indicates the end of the calibration period and start of the forecasting period. The mean (solid red line) and 95% PIs (dashed red lines) of the model fit and forecast are shown.

S8 Fig: COVID-19 death forecasts using daily deaths, sub-epidemic wave model, Mexico City: 30-days ahead forecasts based on the sub-epidemic wave model calibrated using an increasing amount of daily death data (blue circles): 107, 114, 120, 128, 136, 151, 156, 164, 172, 179, 185, 193, 193 epidemic days. The vertical dashed line indicates the end of the calibration period and start of the forecasting period. The mean (solid red line) and 95% PIs (dashed red lines) of the model fit and forecast are shown.

S9 Fig: COVID-19 deaths forecasts using cumulative deaths, GLM model, Mexico: 30-days ahead forecasts based on the Generalized Logistic Growth Model (GLM) calibrated using an increasing amount of cumulative death data (blue circles). The vertical dashed line indicates the end of the calibration period and start of the forecasting period. The mean (solid red line) and 95% PIs (dashed red lines) of the model fit and forecast are shown.

S10 Fig: COVID-19 death forecasts using cumulative deaths, IHME model, Mexico: 30-day ahead forecasts based on the IHME model calibrated using cumulative death data (blue circles). The vertical dashed line indicates the end of the calibration period and start of the forecasting period. The mean (solid red line) and 95% PIs (dashed red lines) of the model fit and forecast are shown.

S11 Fig: COVID-19 death forecasts using cumulative deaths, Richards model, Mexico: 30-day ahead forecasts based on the Richards model calibrated using cumulative death data (blue circles). The vertical dashed line indicates the end of the calibration period and start of the forecasting period. The mean (solid red line) and 95% PIs (dashed red lines) of the model fit and forecast are shown.

S12 Fig: COVID-19 death forecasts using cumulative deaths, sub-epidemic wave model, Mexico: 30-day ahead forecasts based on the Sub-epidemic wave model calibrated using cumulative death data (blue circles). The vertical dashed line indicates the end of the calibration period and start of the forecasting period. The mean (solid red line) and 95% PIs (dashed red lines) of the model fit and forecast are shown.

S13 Fig: COVID-19 deaths forecasts using cumulative deaths, GLM model, Mexico City: 30-day ahead forecasts based on the Generalized Logistic Growth Model (GLM) calibrated using cumulative death data (blue circles). The vertical dashed line indicates the end of the calibration period and start of the forecasting period. The mean (solid red line) and 95% PIs (dashed red lines) of the model fit and forecast are shown.

S14 Fig: COVID-19 death forecasts using cumulative deaths, IHME model, Mexico City: 30-day ahead forecasts based on the IHME model calibrated using cumulative death data (blue circles). The vertical dashed line indicates the end of the calibration period and start of the forecasting period. The mean (solid red line) and 95% PIs (dashed red lines) of the model fit and forecast are shown.

S15 Fig: COVID-19 death forecasts using cumulative deaths, Richards model, Mexico City: 30-day ahead forecasts based on the Richards model calibrated using cumulative death data (blue circles). The vertical dashed line indicates the end of the calibration period and start of the forecasting period. The mean (solid red line) and 95% PIs (dashed red lines) of the model fit and forecast are shown.

S16 Fig: COVID-19 death forecasts using cumulative deaths, sub-epidemic wave model, Mexico City: 30-day ahead forecasts based on the Sub-epidemic wave model calibrated using cumulative death data (blue circles). The vertical dashed line indicates the end of the calibration period and start of the forecasting period. The mean (solid red line) and 95% PIs (dashed red lines) of the model fit and forecast are shown.

S17 Fig: Pre-processing COVID-19 data into incidence rate functions. From left to right: original lab-confirmed COVID-19 cases, curve of daily new cases, smoothed and scaled rate curves, average of rate curves before scaling and smothing.

S18 Fig: Clustering of states according to the shapes of their rate curves. The largest cluster – Cluster 1 – is shown in green while the smallest cluster – Cluster 3 – is shown in the black. One can see that states with similar shapes of rates curves are geographically close to each other.

S19 Fig: Average shapes of the COVID-19 incidence rate curves, along with a one standard-deviation band around the average, in each of the clusters.

S20 Fig: Cluster averages and overall average. These averages represent the four dominant patterns of incidence rates observed across all states.

S21 Fig: Total number of COVID-19 cases as of December 5, 2020

S22: COVID-19 epi-curve overlaid by the curve of stay-at-home orders tweets. Blue line indicates the number of cases by dates of onset and the orange line indicates the number of tweets referring to the stay-at-home orders.

