## Supplemental file for "Transmission dynamics and forecasts of the COVID-19 pandemic in Mexico, March 20-November 11, 2020"

### Model descriptions

#### (i) Generalized logistic growth model

The generalized logistic growth model (GLM) [1] relies on three parameters and allows to capture a range of epidemic growth profiles including polynomial and exponential growth. GLM characterizes epidemic growth by estimating (i) the intrinsic growth rate, $r$ (ii) a dimensionless "deceleration of growth" parameter, $p$ and (iii) $k_0$, representing the final death count. The deceleration parameter modulates the epidemic growth patterns including the exponential growth dynamics (p=1), sub-exponential growth (0<p<1) and constant incidence (p=0). The GLM model is given by the following differential equation:

$$\frac{dC(t)}{dt} = rC(t)^p(1 - \frac{C(t)}{k_0})$$

Where $\frac{dC(t)}{dt}$ describes the deaths over time t. The cumulative number of deaths at time $t$ is given by $C(t)$ while $r$ is a positive parameter denoting the growth rate (1/time), $p\in[0,1]$ is a "deceleration of growth" parameter and $k_0$ is the final number of deaths [1].

#### (ii) Richards growth model

The Richards model [2] also relies on three parameters and extends the simple logistic growth model by incorporating a scaling parameter, *a,* that measures the deviation from the symmetric simple logistic growth curve [2-4]. The Richards model is given by the differential equation:

$$\frac{dC(t)}{dt} = rC(t)\left[1 - \left(\frac{C(t)}{k_o}\right)^a\right]$$

where $C(t)$ represents the cumulative death count at time $t$ , $r$ is the growth rate, $a$ is a scaling parameter and $k_o$ is the final death count.

**(iii) Sub-epidemic wave model.**

We also fit the sub-epidemic model [5] to the daily confirmed COVID -19 death curve, that depicts various profiles of overlapping sub-epidemics shaping the epidemic waves. This model characterizes each group sub-epidemic by a 3-parameter generalized logistic growth model as explained above and given by the following differential equation:

$$\frac{dC(t)}{dt} = rC^{p}(t)\left(1 - \frac{C(t)}{k_o}\right)$$

Next, we model an epidemic wave comprising of *n* overlapping sub-epidemics given by the following system of coupled differential equation:

$$\frac{dC_i(t)}{dt} = rA_{i-1}(t)C_i(t)^p\left(1 - \frac{C_i(t)}{k_i}\right)$$

In this equation $C_i(t)$ describes the cumulative death number for i[th] sub-epidemic, and $k_i$ is the size of sub-epidemic i where i=1….n. $A_i(t)$ is an indicator variable that models the onset timing of (i+1)th sub-epidemic, making sure that sub-epidemics comprising an epidemic wave follow a regular structure. Therefore,

$$A_i(t) = \begin{cases} 1 & C_i(t) > C_{thr} \\ 0 & Otherwise \end{cases} \quad i = 1,2,3,\dots n$$

Where $1 \leq C_{thr} < k_o$ and $A_1(t) = 1$ for the sub-epidemic 1. Moreover, for the subsequently occurring sub-epidemics, the size of i[th] sub-epidemic ($k_i$) declines exponentially at a rate q. This occurs owing to multiple factors including the behavior changes, effect of interventions, and changes in disease transmission dependent on seasonality. If q=0, then the sub-epidemic model predicts an epidemic wave composed of equal sized sub-epidemics [5]. If we assume that the subsequent sub-epidemic sizes decline exponentially, we get

$$k_i = k_0 e^{-q(i-1)}$$

Where $k_0$ is the final size of the epidemic (when the epidemic ends). Hence, if the epidemic wave is comprised by a single sub-epidemic, the sub-epidemic model shrinks to the three-parameter generalized growth model whereas an epidemic wave comprised by two or more sub-epidemics is calibrated with five parameters: $r, p, k_o, q$ *and* $C_{thr}$.

**(iv) IHME model**

We compare the results of our short term forecasts with the Institute for Health Metrics and Evaluation (IHME) model briefly described in ref [6]. This model utilized death data from a range of both governmental, non-profit, and volunteer organizations by the date of reporting. Observed cumulative deaths were smoothed via spline based smoothing algorithm with randomly placed knots. Resampling and bootstrapping of observed deaths was performed to introduce uncertainty in the data. The time series of case data was used as an indicator of death based on infection fatality ratio (IFR) and a lag from COVID-19 infection to death. Estimated infections based on an age-distribution of infections and on age-specific IFRs were derived from the smoothed estimates of observed deaths by location. Lastly, the age-specific infections were merged into total infections by day and state and used as data inputs in the SEIR model [6].

#### (v) Generalized growth model

Generalized growth model (GGM) characterizes the early ascending phase of the epidemic by estimating two parameters: (1) the intrinsic growth rate, $r$; and (2) a dimensionless “deceleration of growth” parameter, $p$. This model allows to capture a range of epidemic growth profiles by modulating the deceleration of growth parameter, $p$. The GGM model is given by the following differential equation:

$$\frac{dC(t)}{dt} = C'(t) = rC(t)^p$$

In this equation $C'(t)$ describes the incidence curve over time $t$, solution $C(t)$ describes the cumulative number of cases at time $t$ and $p \in [0,1]$ is a “deceleration of growth” parameter. This equation depicts constant incidence over time if $p$=0 and becomes an exponential growth model for cumulative cases if $p$ =1. Whereas if $p$ is in the range 0< $p$ <1, then the model indicates sub-exponential growth dynamics [3, 7].

### Figures

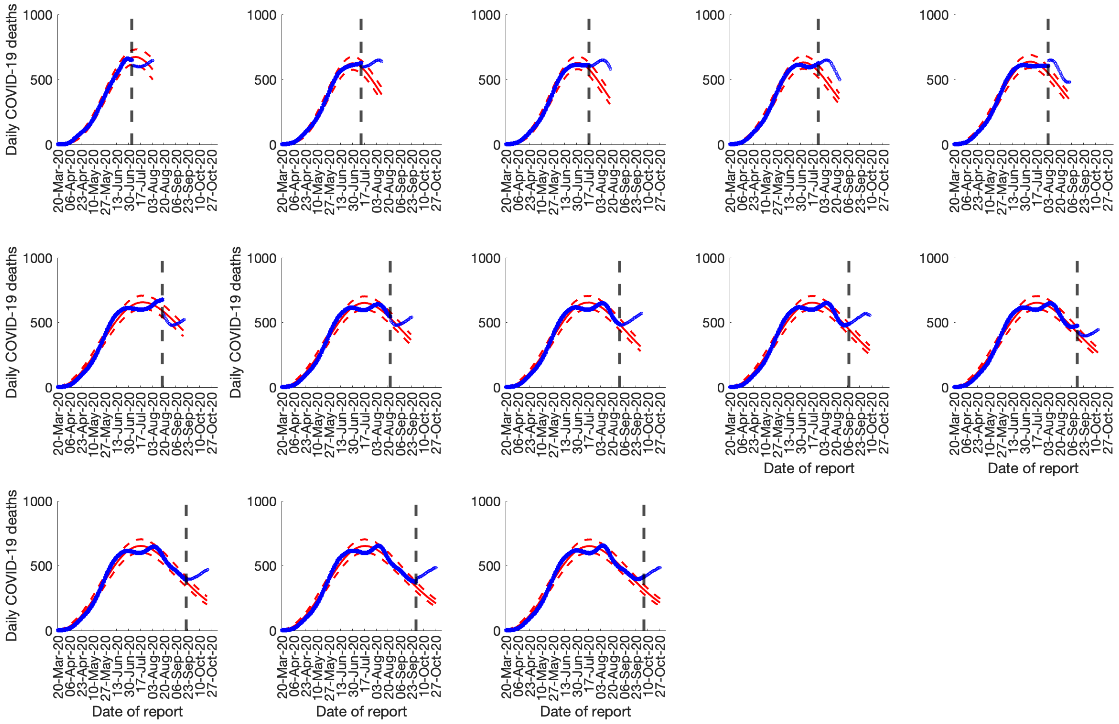


S1 Fig: COVID-19 deaths forecasts using daily deaths, GLM model, Mexico: 30-days ahead forecasts based on the Generalized Logistic Growth Model (GLM) calibrated using an increasing amount of daily death data (blue circles): 107, 114, 120, 128, 136, 151, 156, 164, 172, 179, 185, 193, 193 epidemic days. The vertical dashed line indicates the end of the calibration period and start of the forecasting period. The mean (solid red line) and 95% PIs (dashed red lines) of the model fit and forecast are shown.

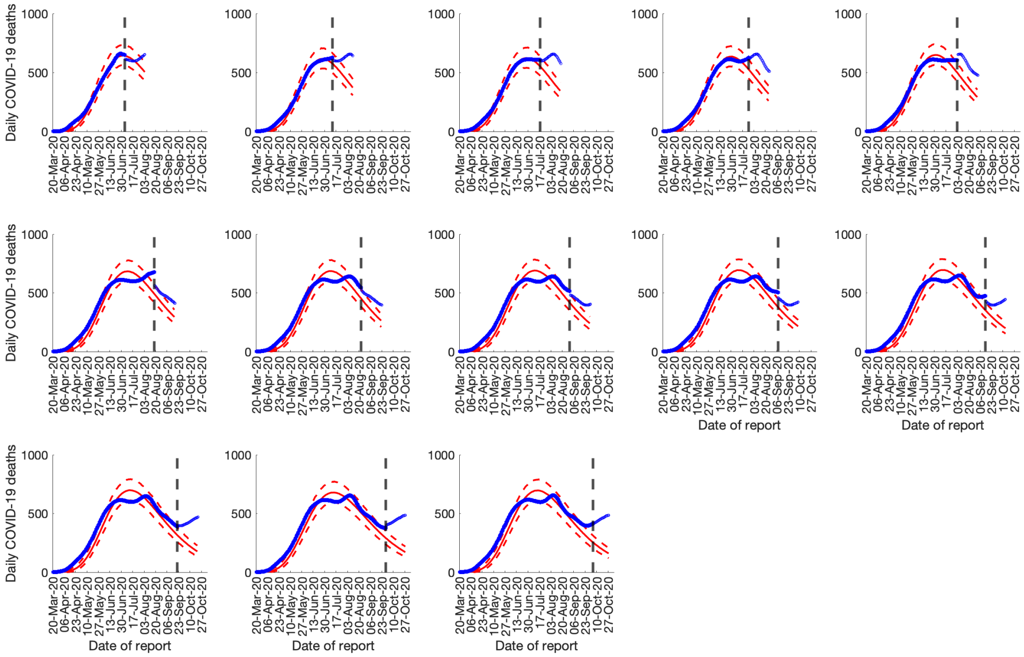


S2 Fig: COVID-19 death forecasts using daily deaths, Richards model, Mexico: 30-days ahead forecasts based on the Richards model calibrated using an increasing amount of daily death data (blue circles): 107, 114, 120, 128, 136, 151, 156, 164, 172, 179, 185, 193, 193 epidemic days. The vertical dashed line indicates the end of the calibration period and start of the forecasting period. The mean (solid red line) and 95% PIs (dashed red lines) of the model fit and forecast are shown

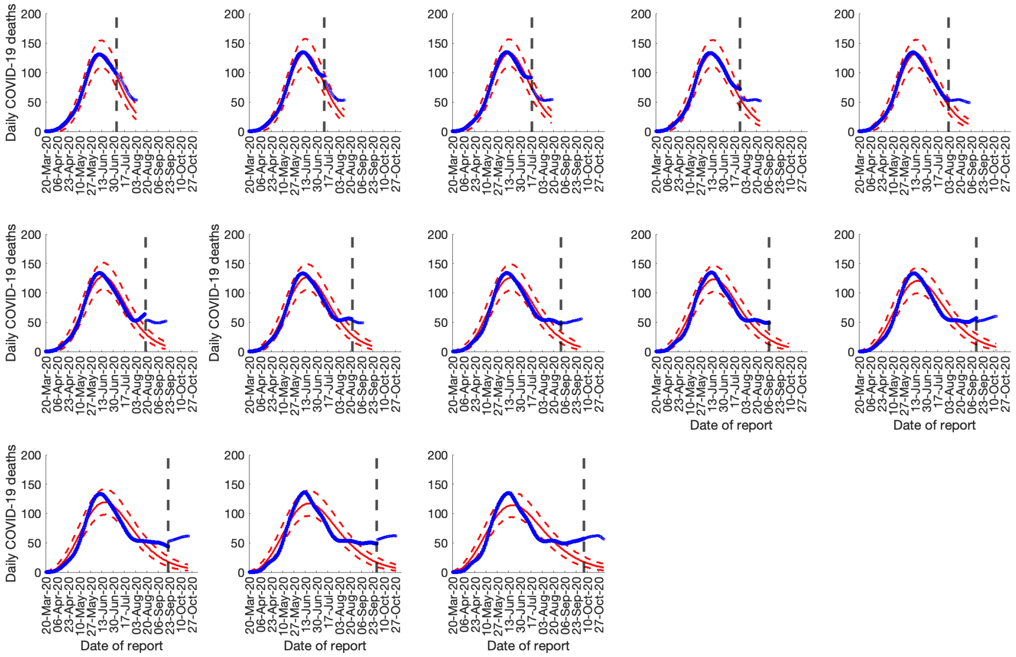


S3 Fig: COVID-19 death forecasts using daily deaths, GLM model, Mexico City: 30-days ahead forecasts based on the GLM model calibrated using an increasing amount of daily death data (blue circles): 107, 114, 120, 128, 136, 151, 156, 164, 172, 179, 185, 193, 193 epidemic days. The vertical dashed line indicates the end of the calibration period and start of the forecasting period. The mean (solid red line) and 95% PIs (dashed red lines) of the model fit and forecast are shown.

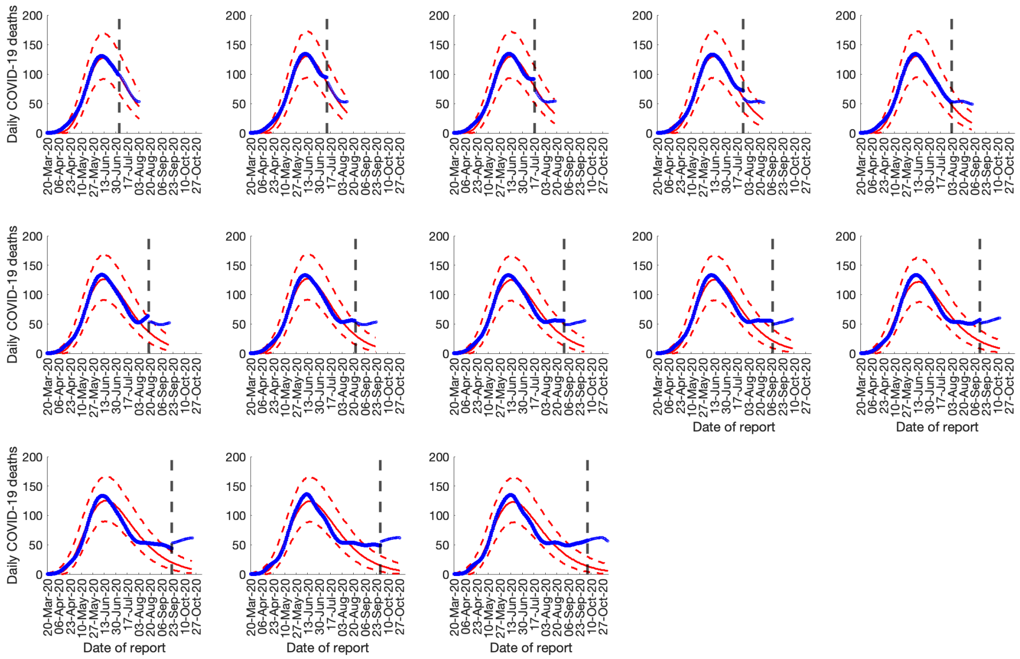


S4 Fig: COVID-19 death forecasts using daily deaths, Richards model, Mexico City: 30-days ahead forecasts based on the Richards model calibrated using an increasing amount of daily death data (blue circles): 107, 114, 120, 128, 136, 151, 156, 164, 172, 179, 185, 193, 193epidemic days. The vertical dashed line indicates the end of the calibration period and start of the forecasting period. The mean (solid red line) and 95% PIs (dashed red lines) of the model fit and forecast are shown.

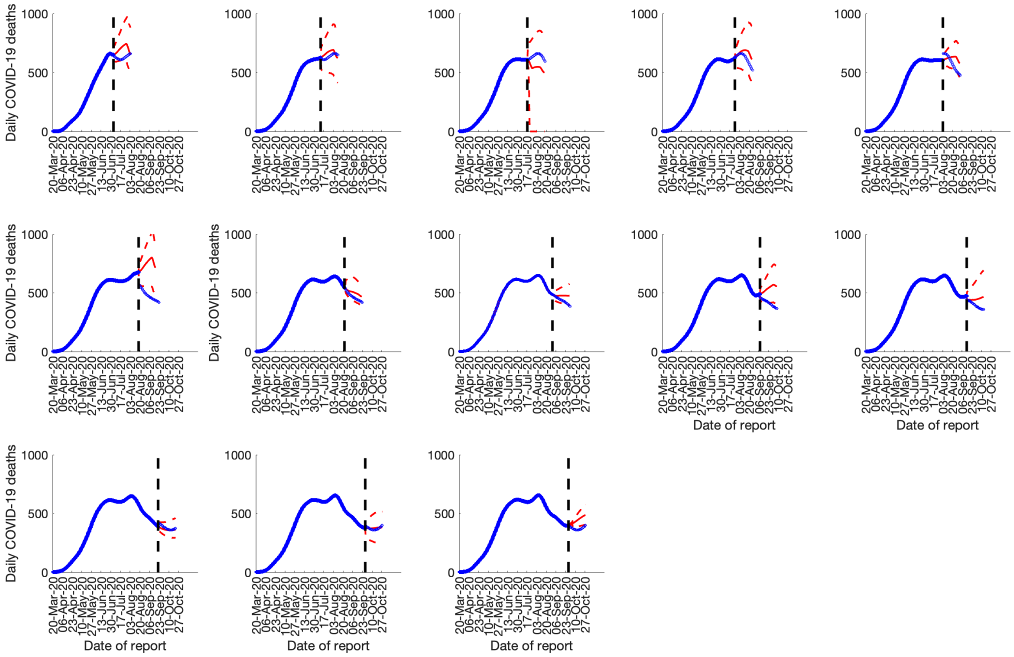


S5 Fig: COVID-19 death forecasts using daily deaths, IHME model, Mexico: 30-days ahead forecasts based on the IHME model calibrated using an increasing amount of daily death data (blue circles): 107, 114, 120, 128, 136, 151, 156, 164, 172, 179, 185, 193, 193 epidemic days. The vertical dashed line indicates the end of the calibration period and start of the forecasting period. The mean (solid red line) and 95% PIs (dashed red lines) of the model fit and forecast are shown.

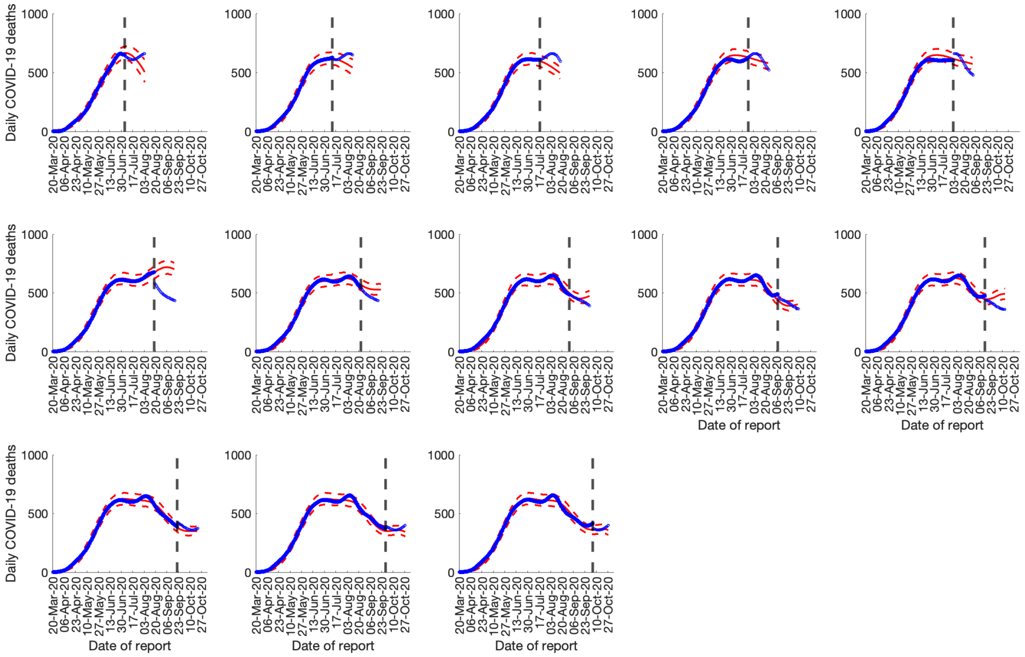


S6 Fig: COVID-19 death forecasts using daily deaths, sub-epidemic wave model, Mexico: 30-days ahead forecasts based on the sub-epidemic wave model calibrated using an increasing amount of daily death data (blue circles): 107, 114, 120, 128, 136, 151, 156, 164, 172, 179, 185, 193, 193 epidemic days. The vertical dashed line indicates the end of the calibration period and start of the forecasting period. The mean (solid red line) and 95% PIs (dashed red lines) of the model fit and forecast are shown.

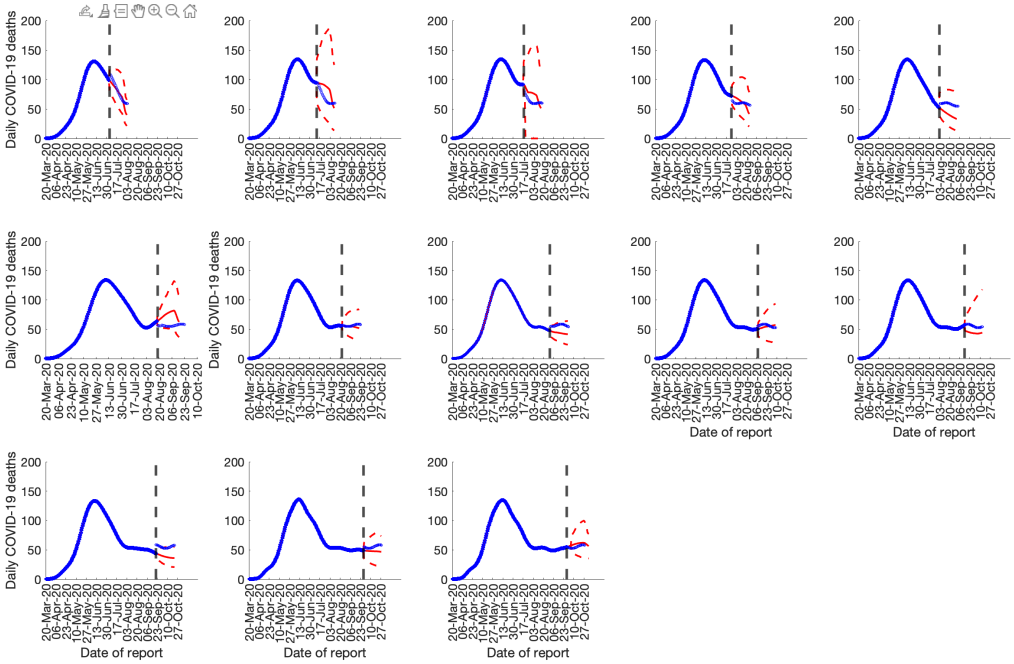


S7 Fig: COVID-19 death forecasts using daily deaths, IHME model, Mexico City: 30-days ahead forecasts based on the IHME model calibrated using an increasing amount of daily death data (blue circles): 107, 114, 120, 128, 136, 151, 156, 164, 172, 179, 185, 193, 193 epidemic days. The vertical dashed line indicates the end of the calibration period and start of the forecasting period. The mean (solid red line) and 95% PIs (dashed red lines) of the model fit and forecast are shown.

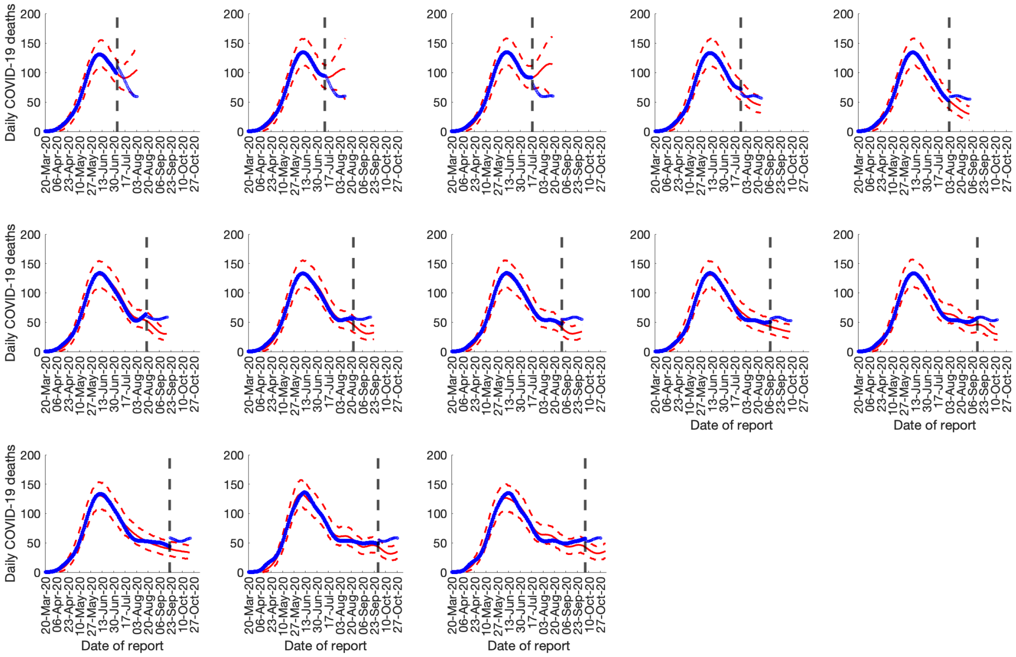


S8 Fig: COVID-19 death forecasts using daily deaths, sub-epidemic wave model, Mexico City: 30-days ahead forecasts based on the sub-epidemic wave model calibrated using an increasing amount of daily death data (blue circles): 107, 114, 120, 128, 136, 151, 156, 164, 172, 179, 185, 193, 193 epidemic days. The vertical dashed line indicates the end of the calibration period and start of the forecasting period. The mean (solid red line) and 95% PIs (dashed red lines) of the model fit and forecast are shown.

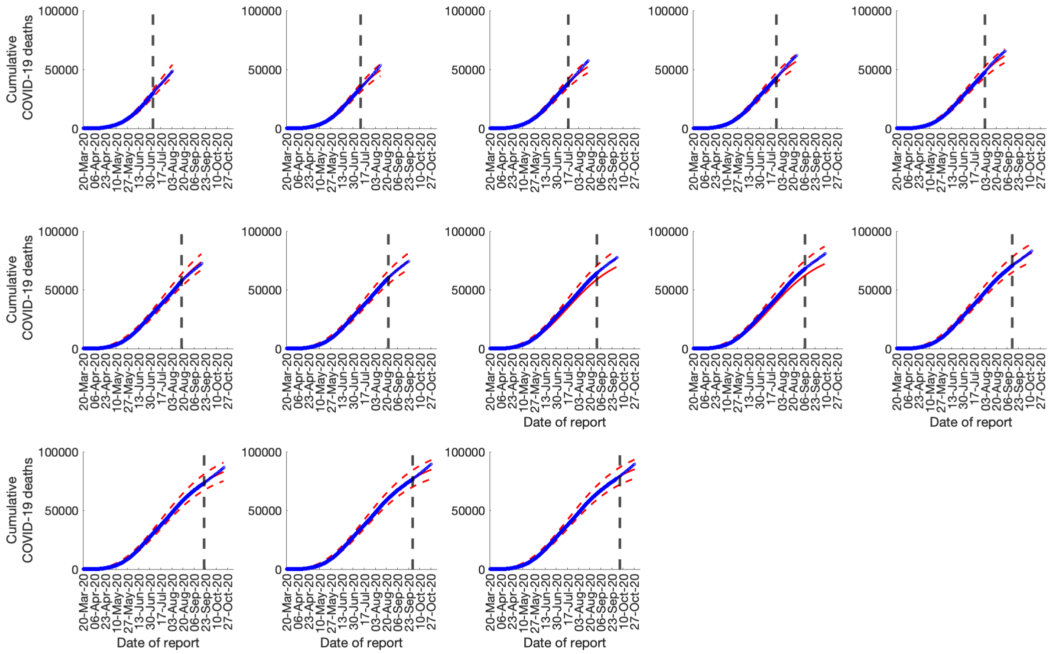


S9 Fig: COVID-19 deaths forecasts using cumulative deaths, GLM model, Mexico: 30-days ahead forecasts based on the Generalized Logistic Growth Model (GLM) calibrated using an increasing amount of cumulative death data (blue circles). The vertical dashed line indicates the end of the calibration period and start of the forecasting period. The mean (solid red line) and 95% PIs (dashed red lines) of the model fit and forecast are shown.

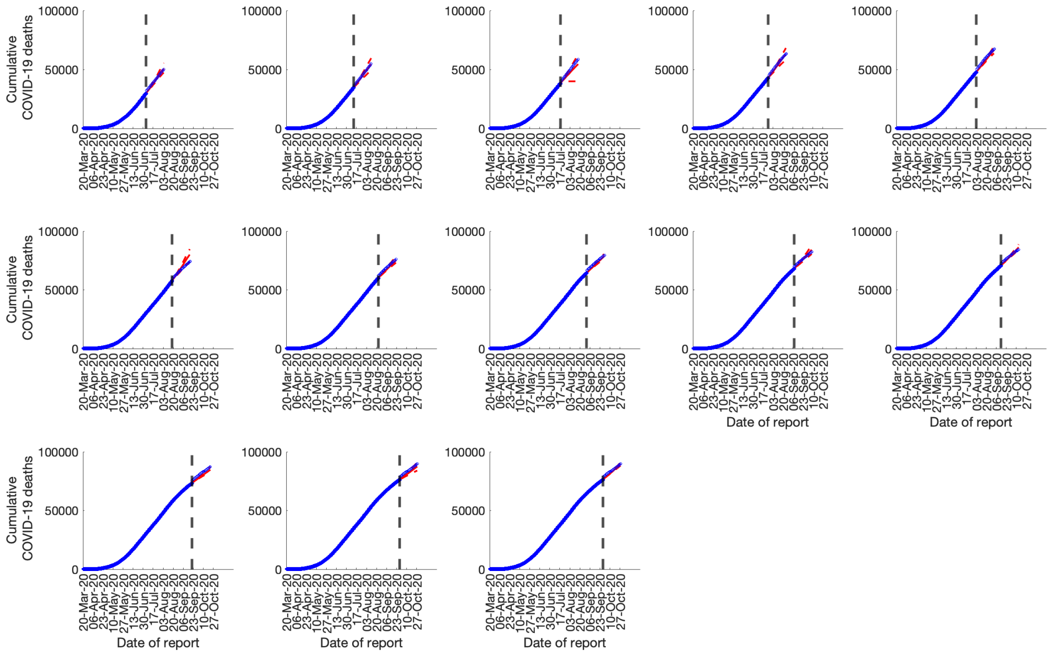


S10 Fig: COVID-19 death forecasts using cumulative deaths, IHME model, Mexico: 30-day ahead forecasts based on the IHME model calibrated using cumulative death data (blue circles). The vertical dashed line indicates the end of the calibration period and start of the forecasting period. The mean (solid red line) and 95% PIs (dashed red lines) of the model fit and forecast are shown.

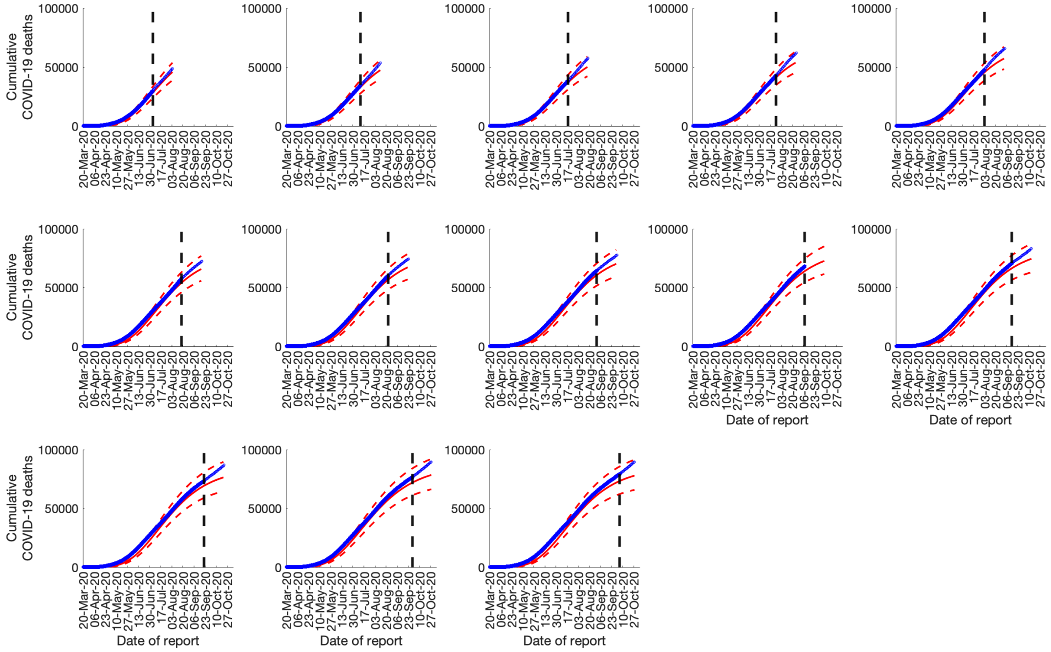


S11 Fig: COVID-19 death forecasts using cumulative deaths, Richards model, Mexico: 30-day ahead forecasts based on the Richards model calibrated using cumulative death data (blue circles). The vertical dashed line indicates the end of the calibration period and start of the forecasting period. The mean (solid red line) and 95% PIs (dashed red lines) of the model fit and forecast are shown.

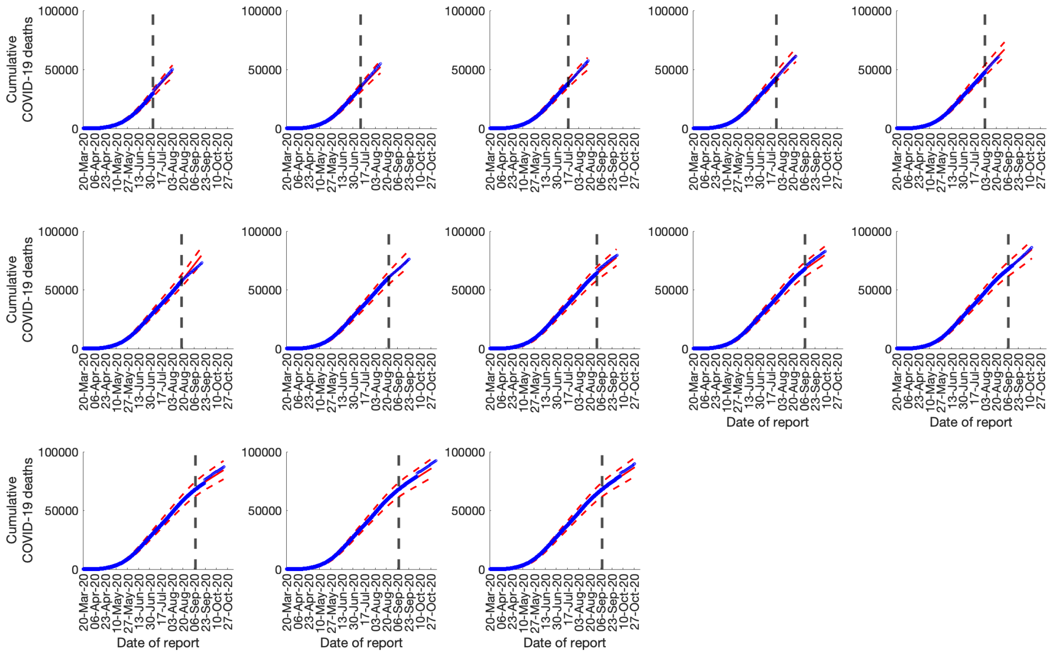


S12 Fig: COVID-19 death forecasts using cumulative deaths, sub-epidemic wave model, Mexico: 30-day ahead forecasts based on the sub-epidemic wave model calibrated using cumulative death data (blue circles). The vertical dashed line indicates the end of the calibration period and start of the forecasting period. The mean (solid red line) and 95% PIs (dashed red lines) of the model fit and forecast are shown.

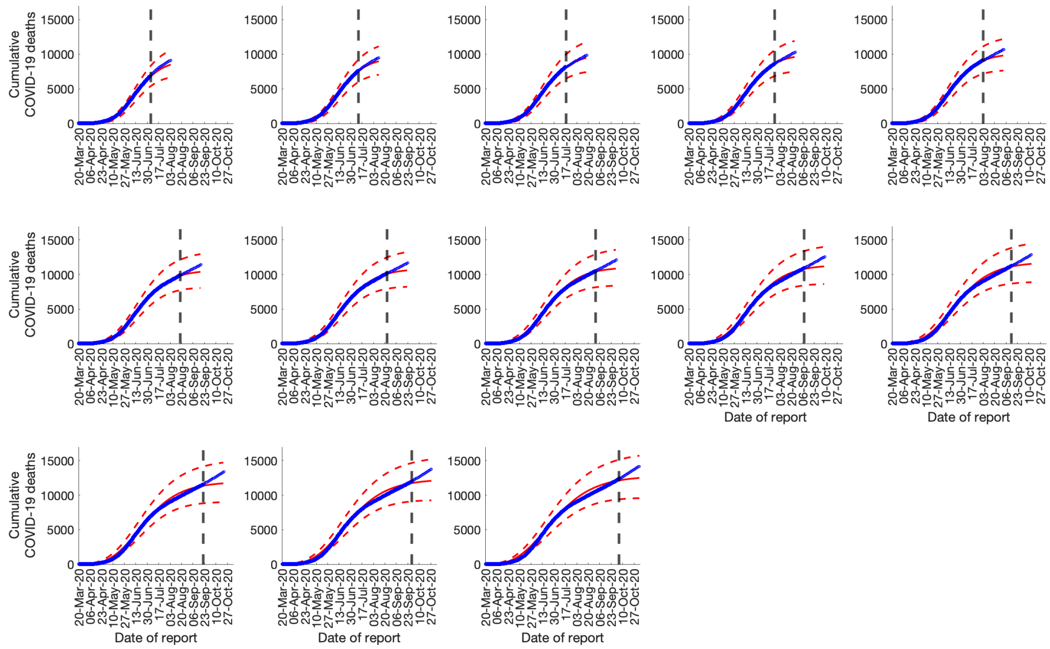


S13 Fig: COVID-19 deaths forecasts using cumulative deaths, GLM model, Mexico City: 30-day ahead forecasts based on the Generalized Logistic Growth Model (GLM) calibrated using cumulative death data (blue circles). The vertical dashed line indicates the end of the calibration period and start of the forecasting period. The mean (solid red line) and 95% PIs (dashed red lines) of the model fit and forecast are shown.

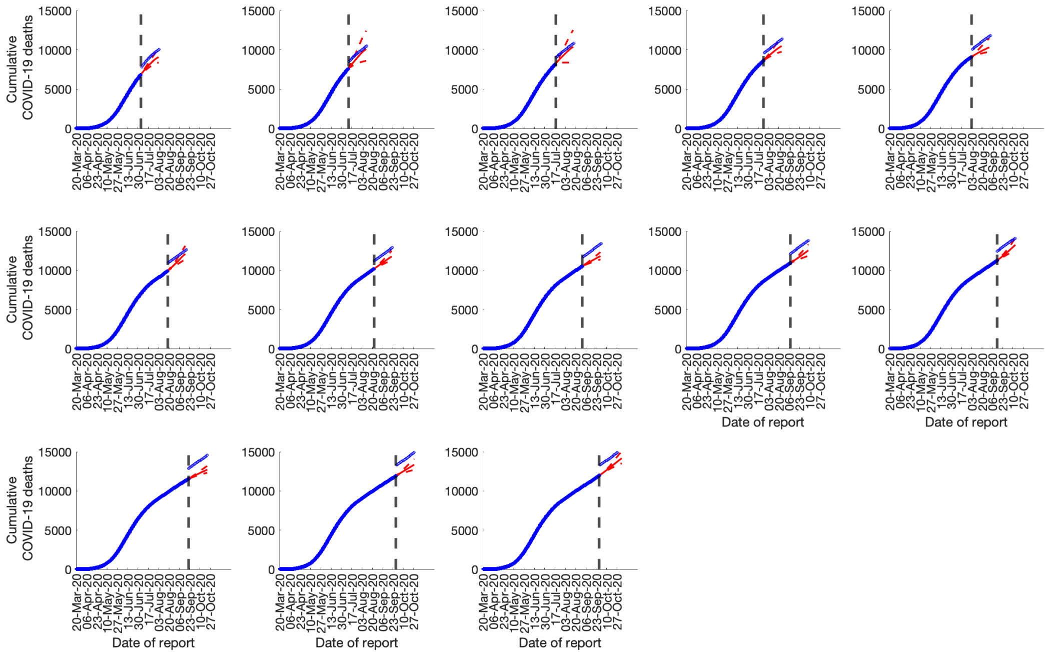


S14 Fig: COVID-19 death forecasts using cumulative deaths, IHME model, Mexico City: 30-day ahead forecasts based on the IHME model calibrated using cumulative death data (blue circles). The vertical dashed line indicates the end of the calibration period and start of the forecasting period. The mean (solid red line) and 95% PIs (dashed red lines) of the model fit and forecast are shown.

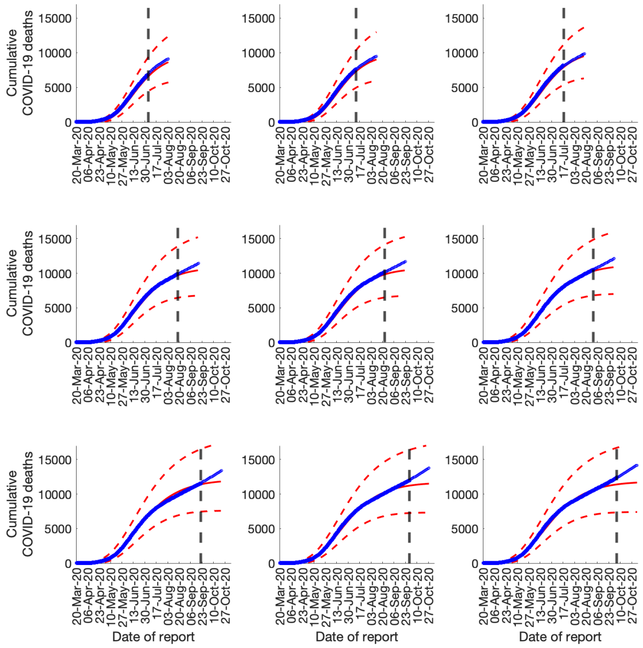


S15 Fig: COVID-19 death forecasts using cumulative deaths, Richards model, Mexico City: 30-day ahead forecasts based on the Richards model calibrated using cumulative death data (blue circles). The vertical dashed line indicates the end of the calibration period and start of the forecasting period. The mean (solid red line) and 95% PIs (dashed red lines) of the model fit and forecast are shown.

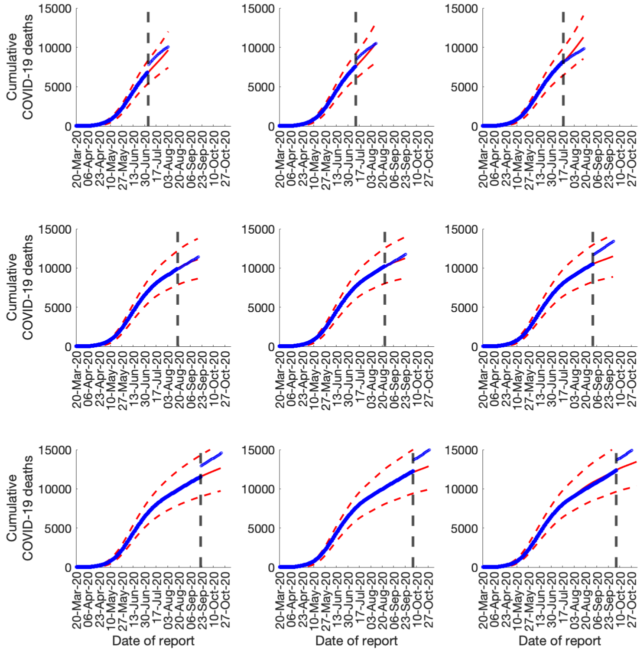


S16 Fig: COVID-19 death forecasts using cumulative deaths, sub-epidemic wave model, Mexico City: 30-day ahead forecasts based on the sub-epidemic wave model calibrated using cumulative death data (blue circles). The vertical dashed line indicates the end of the calibration period and start of the forecasting period. The mean (solid red line) and 95% PIs (dashed red lines) of the model fit and forecast are shown.

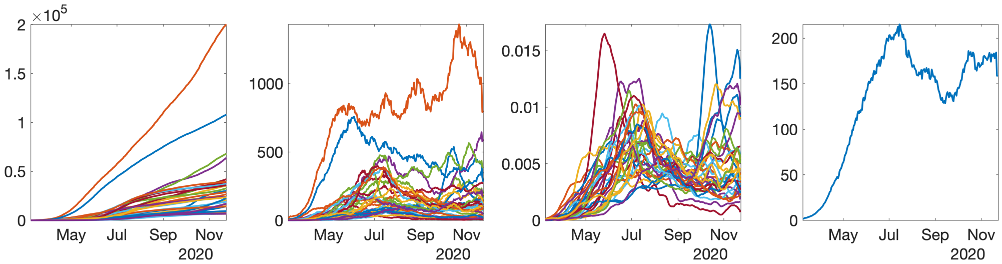


S17 Fig: Pre-processing COVID-19 data into incidence rate functions. From left to right: original lab-confirmed COVID-19 cases, curve of daily new cases, smoothed and scaled rate curves, average of rate curves before scaling and smothing.

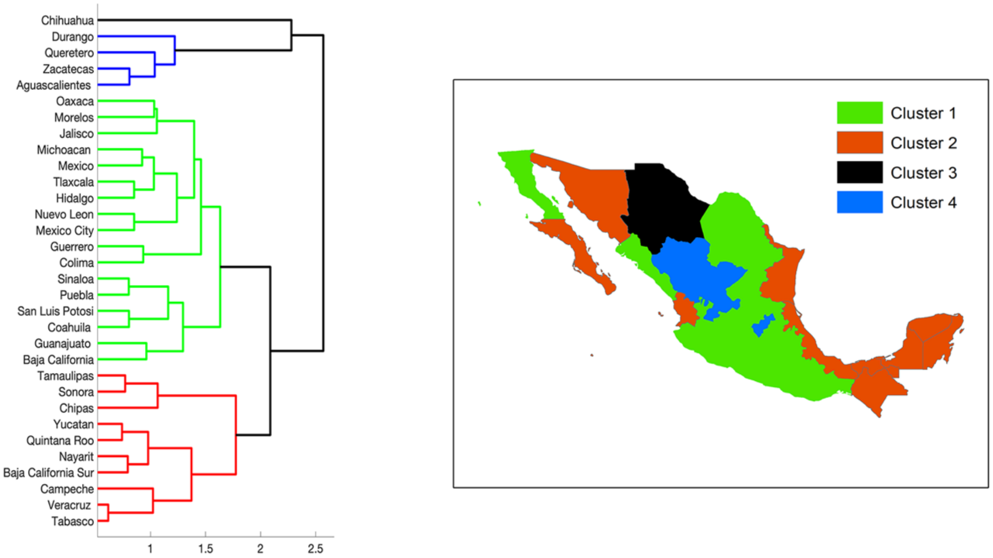


S18 Fig: Clustering of states according to the shapes of their rate curves. The largest cluster – Cluster 1 – is shown in green while the smallest cluster – Cluster 3 – is shown in the black. One can see that states with similar shapes of rates curves are geographically close to each other.

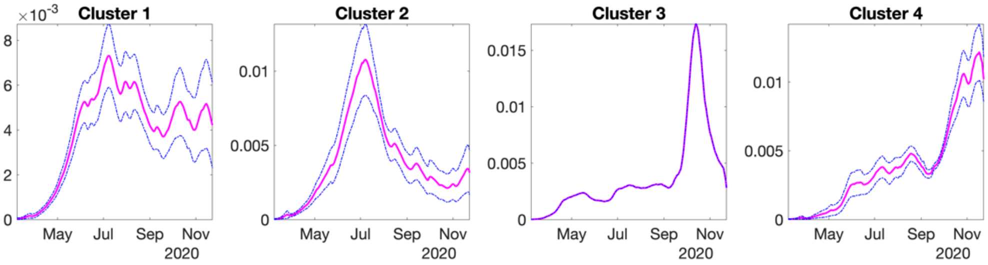


S19 Fig: Average shapes of the COVID-19 incidence rate curves, along with a one standard-deviation band around the average, in each of the clusters.

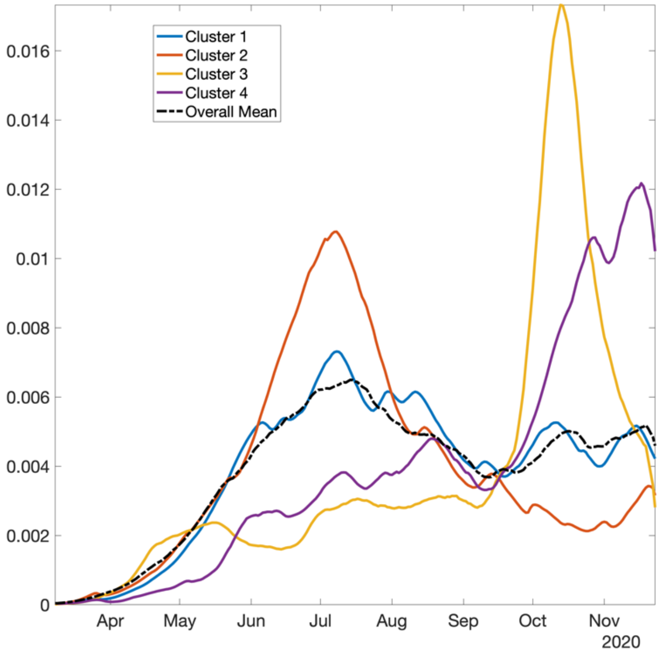


S20 Fig: Cluster averages and overall average. These averages represent the four dominant patterns of incidence rates observed across all states.

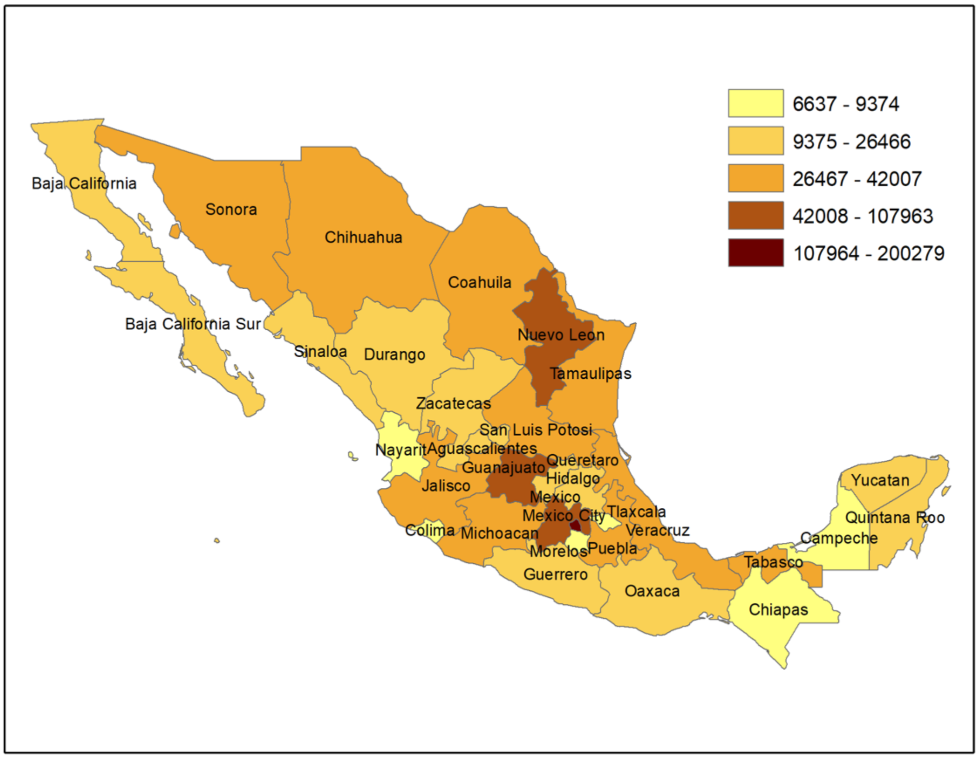


S21 Fig: Total number of COVID-19 cases as of December 5, 2020

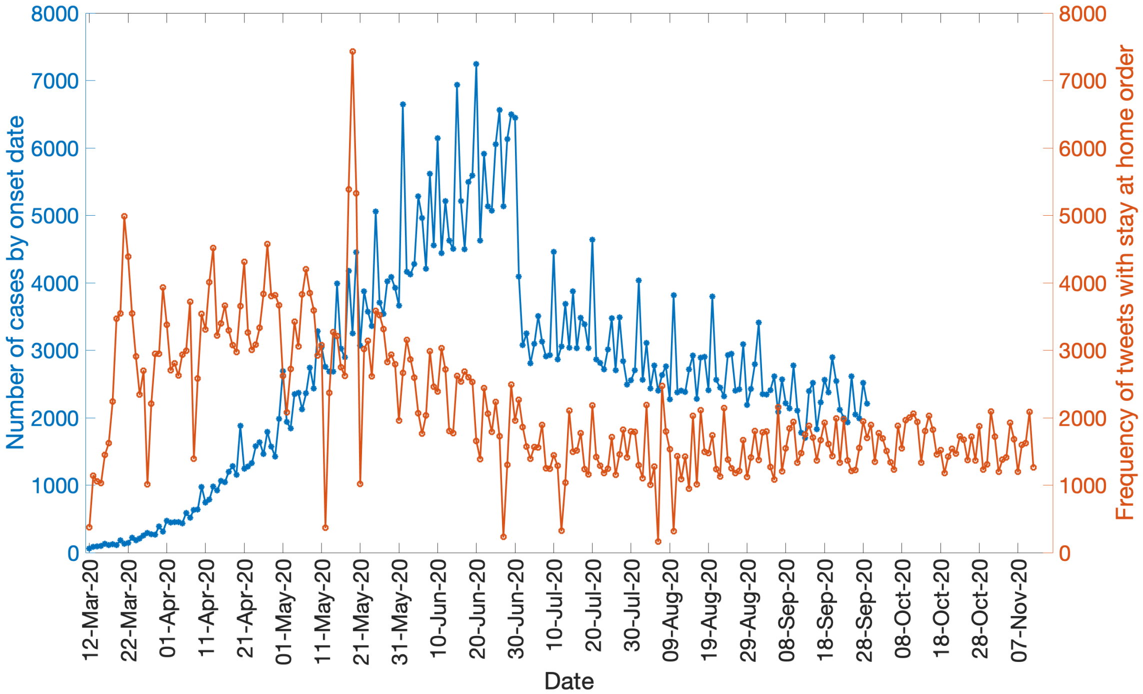


S22 Fig: COVID-19 epi-curve overlaid by the curve of stay-at-home orders tweets. Blue line indicates the number of cases by dates of onset and the orange line indicates the number of tweets referring to the stay at home orders.

Table S1: Calibration performance metrics for the country of Mexico as of November 11, 2020. Higher 95% PI coverage and lower RMSE, MAE and MIS represent better performance.

| Calibration period | Number of days for calibration period | RMSE | MAE | MIS | PI |
|---|---|---|---|---|---|
| GLM | | | | | |
| 3/20-9/27 | 193 | 2.9048 | 23.3109 | 132.4455 | 70.83 |
| 3/20-9/27 | 193 | **2.7899**[a] | 21.6382 | 120.2581 | 70.31 |
| 3/20-9/20 | 185 | 2.8006 | 21.5245 | 111.5216 | 69.19 |
| 3/20-9/13 | 179 | 3.0168 | 24.5451 | 132.1344 | 66.48 |
| 3/20-9/7 | 172 | **2.9599**[a] | 24.1823 | 111.6296 | 68.86 |
| 3/20-8/30 | 164 | **2.9532**[a] | 23.2876 | **103.2752**[a] | 71.3 |
| 3/20-8/22 | 156 | 3.251 | 24.1819 | 112.0674 | 64.74 |
| 3/20-8/17 | 151 | 3.8743 | 29.5499 | 206.0324 | 47.68 |
| 3/20-8/2 | 136 | **1.8125**[a] | 15.1316 | 69.8466 | 97.79 |
| 3/20-7/25 | 128 | **1.0388**[a] | **11.5729**[a] | 70.8107 | **97.66**[a] |
| 3/20-7/17 | 120 | 0.0598 | 7.2138 | **60.6835**[a] | **100**[a] |
| 3/20-7/11 | 114 | **0.1863**[a] | 7.2144 | **59.0629**[a] | **100**[a] |
| 3/20-7/4 | 107 | 1.07514 | **5.2267**[a] | **57.8046**[a] | **98.13**[a] |
| Richards model | | | | | |
| 3/20-9/27 | 192 | 24.9399 | 51.5158 | 579.2161 | 52.6 |
| 3/20-9/27 | 192 | 22.3437 | 43.0289 | 549.6926 | 55.73 |
| 3/20-9/20 | 185 | 21.7383 | 50.4173 | 509.0301 | 55.14 |
| 3/20-9/13 | 178 | 22.9219 | 51.6033 | 552.0598 | 51.69 |
| 3/20-9/7 | 172 | 21.9109 | 50.9341 | 513.151 | 54.07 |
| 3/20-8/30 | 164 | 20.2246 | 49.1298 | 420.1666 | 57.32 |
| 3/20-8/22 | 156 | 20.9625 | 49.9921 | 472.6152 | 50 |
| 3/20-8/17 | 151 | 21.0673 | 51.995 | 571.9809 | 52.32 |
| 3/20-8/2 | 136 | 13.4568 | 27.7342 | 252.2454 | 73.53 |
| 3/20-7/25 | 128 | 13.5594 | 22.6754 | 250.4846 | 74.22 |
| 3/20-7/17 | 120 | 11.7392 | 18.0134 | 228.4447 | 75 |
| 3/20-7/11 | 114 | 10.2078 | 14.8153 | 194.3025 | 74.56 |
| 3/20-7/4 | 107 | 13.74 | 22.8536 | 312.8941 | 71.96 |
| Sub-epidemic model | | | | | |
| 3/20-9/27 | 192 | **2.501**[a] | **12.9893**[a] | **86.104**[a] | **91.67**[a] |
| 3/20-9/27 | 192 | 4.6878 | **11.8312**[a] | **79.4903**[a] | **92.71**[a] |

| | | | | | |
|---|---|---|---|---|---|
| 3/20-9/20 | 185 | **1.6071**[a] | **10.8071**[a] | **72.56**[a] | **99.46**[a] |
| 3/20-9/13 | 178 | **1.5606**[a] | **10.5654**[a] | **91.6648**[a] | **89.33**[a] |
| 3/20-9/7 | 172 | 3.6014 | **11.5299**[a] | **85.3011**[a] | **90.12**[a] |
| 3/20-8/30 | 164 | 2.9803 | **11.2318**[a] | 118.4495 | **84.76**[a] |
| 3/20-8/22 | 156 | **2.1126**[a] | **9.5039**[a] | **81.1916**[a] | **90.38**[a] |
| 3/20-8/17 | 151 | **1.2807**[a] | **7.3038**[a] | **69.4787**[a] | **99.34**[a] |
| 3/20-8/2 | 136 | 10.4909 | **13.7231**[a] | **67.8069**[a] | **99.26**[a] |
| 3/20-7/25 | 128 | 9.8848 | 13.0077 | **66.9836**[a] | 96.09 |
| 3/20-7/17 | 120 | **0.0343**[a] | **4.2524**[a] | 62.2455 | 99.17 |
| 3/20-7/11 | 114 | 0.7179 | **5.5247**[a] | 60.85 | 99.12 |
| 3/20-7/4 | 107 | **0.9118**[a] | 5.3033 | 61.9557 | 88.79 |

[a]Best performance with regards to the performance metric (row) per calibration period, i.e. highest prediction interval (PI) coverage and lowest mean interval score (MIS), mean squared error (MSE) and mean absolute error (MAE).

Table S2: Calibration performance metrics for the Mexico City as of November 11, 2020. Higher 95% PI coverage and lower RMSE, MAE and MIS represent better performance.

| Calibration period | Number of calibration days | RMSE | MAE | MIS | PI |
|---|---|---|---|---|---|
| GLM | | | | | |
| 3/20-9/27 | 199 | 0.915 | 13.9516 | 170.137 | 53.27 |
| 3/20-9/27 | 193 | 0.8116 | 12.5158 | 133.8038 | 63.02 |
| 3/20-9/20 | 185 | 0.6951 | 11.2254 | 102.4674 | 73.51 |
| 3/20-9/13 | 178 | 0.6612 | 10.7864 | 102.8431 | 77.65 |
| 3/20-9/7 | 172 | 0.5033 | 9.1957 | 80.1844 | 83.72 |
| 3/20-8/30 | 164 | 0.3498 | 7.7533 | 64.7135 | 89.63 |
| 3/20-8/22 | 157 | 0.2209 | 6.8029 | 62.3701 | 91.72 |
| 3/20-8/17 | 151 | **0.1658**[a] | 5.8284 | 58.8222 | 94.04 |
| 3/20-8/2 | 136 | 0.0197 | 3.0926 | **28.0961**[a] | **100**[a] |
| 3/20-7/25 | 128 | **0.0306**[a] | 3.1719 | 29.1193 | 99.92 |
| 3/20-7/17 | 120 | 0.2245 | 3.2924 | **28.8139**[a] | 99.17 |
| 3/20-7/11 | 114 | 0.1678 | 2.6467 | **27.0702**[a] | **100**[a] |
| 3/20-7/4 | 107 | 0.3373 | 1.942 | **26.4446**[a] | **100**[a] |
| Richards model | | | | | |
| 3/20-9/27 | 199 | 4.0688 | 11.4509 | 129.2115 | 85.93 |
| 3/20-9/27 | 192 | 3.0002 | 10.1032 | 93.2947 | 88.54 |
| 3/20-9/20 | 185 | 0.1518 | 8.9106 | 61.6089 | 91.89 |
| 3/20-9/13 | 178 | 2.6208 | 8.1841 | 72.8783 | 93.26 |
| 3/20-9/7 | 172 | 2.1165 | 7.0608 | 58.8552 | 94.77 |
| 3/20-8/30 | 164 | 0.4029 | 6.2239 | 48.071 | 99.39 |
| 3/20-8/22 | 156 | 1.2666 | 5.1125 | 49.4908 | 98.08 |
| 3/20-8/17 | 151 | 0.399 | 5.0252 | 49.9194 | 98.68 |
| 3/20-8/2 | 136 | 0.8998 | 2.3949 | 47.2502 | **100**[a] |
| 3/20-7/25 | 128 | 0.9866 | 2.9086 | 47.34 | **100**[a] |
| 3/20-7/17 | 120 | 1.024 | 3.3175 | 47.1177 | **100**[a] |
| 3/20-7/11 | 114 | 1.1465 | 3.0077 | 45.6331 | **100**[a] |
| 3/20-7/4 | 107 | 1.2738 | 2.6535 | 44.4158 | 98.13 |
| Sub-epidemic model | | | | | |
| 3/20-9/27 | 199 | **0.2287**[a] | **4.8811**[a] | **29.2626**[a] | **98.49**[a] |
| 3/20-9/27 | 192 | **0.5046**[a] | **4.1335**[a] | **35.6951**[a] | **91.67**[a] |
| 3/20-9/20 | 185 | **0.1158**[a] | **3.4047**[a] | **28.7305**[a] | **99.46**[a] |
| 3/20-9/13 | 178 | **0.2494**[a] | **3.3685**[a] | **28.7253**[a] | **96.63**[a] |
| 3/20-9/7 | 172 | **0.3476**[a] | **2.8924**[a] | **29.152**[a] | **99.42**[a] |

| | | | | | |
|---|---|---|---|---|---|
| 3/20-8/30 | 164 | **0.0438**[a] | **2.5443**[a] | **28.9252**[a] | **99.39**[a] |
| 3/20-8/22 | 157 | **0.1072**[a] | **2.4802**[a] | **28.9528**[a] | **99.36**[a] |
| 3/20-8/17 | 151 | 0.4338 | **2.8826**[a] | **29.0623**[a] | **99.34**[a] |
| 3/20-8/2 | 136 | **0.00083134**[a] | **1.238**[a] | 29.2342 | 99.26 |
| 3/20-7/25 | 128 | 0.1133 | **1.6842**[a] | **28.9927**[a] | 99.22 |
| 3/20-7/17 | 120 | **0.1815**[a] | **1.0953**[a] | 29.0969 | 99.17 |
| 3/20-7/11 | 114 | **0.0683**[a] | **1.3301**[a] | 28.4282 | 99.12 |
| 3/20-7/4 | 107 | **0.0129**[a] | **1.318**[a] | 27.8169 | 99.07 |

[a]Best performance with regards to the performance metric (row) per calibration period, i.e. highest prediction interval (PI) coverage and lowest mean interval score (MIS), mean squared error (MSE) and mean absolute error (MAE).

Table S3 Forecasting performance metrics for Mexico City as of November 11, 2020. Higher 95% PI coverage and lower RMSE, MAE and MIS represent better performance.

| Forecast period | RMSE | MAE | MIS | PI |
|---|---|---|---|---|
| GLM | | | | |
| 9/28-11/03 | 368.8303 | 368.1496 | 15555.00 | 0 |
| 9/28-10/27 | 362.6612 | 362.4413 | 15068.00 | 0 |
| 9/21-10/21 | 368.3875 | 367.9834 | 15069.00 | 0 |
| 9/14-10/13 | 42.5969 | 42.5609 | 2021.30 | 0 |
| 9/8-10/8 | 43.3936 | 43.3061 | 1879.10 | 0 |
| 8/31-9/29 | 42.9048 | 42.5807 | 1805.60 | 0 |
| 8/23-9/21 | 39.7153 | 38.9318 | 1543.60 | 0 |
| 8/18-9/16 | 38.7439 | 38.3321 | 1475.70 | 0 |
| 8/3-9/1 | 28.3197 | 27.4481 | 824.03 | 3.3 |
| 7/26-8/24 | 28.9153 | 27.0895 | 760.94 | 16.67 |
| 7/18-8/16 | 21.5585 | 19.363 | 436.76 | 46.67 |
| 7/12-8/10 | 22.3979 | 21.6227 | 397.80 | 23.33 |
| 7/5-8/3 | 21.4829 | 21.3684 | 291.25 | 16.67 |
| Richards model | | | | |
| 9/28-11/03 | 46.4734 | 46.298 | 2211.50 | 0 |
| 9/28-10/27 | 44.86 | 44.6312 | 1854.00 | 0 |
| 9/21-10/21 | 41.003 | 40.8629 | 1426.40 | 0 |
| 9/14-10/13 | 41.1357 | 41.1021 | 1475.60 | 0 |
| 9/8-10/8 | 42.7717 | 42.6453 | 1403.00 | 0 |
| 8/31-9/29 | 39.0744 | 38.5684 | 1086.30 | 0 |
| 8/23-9/21 | 37.5132 | 36.9526 | 1.03E+03 | 0 |
| 8/18-9/16 | 29.0641 | 28.8758 | 728.7203 | 5 |
| 8/3-9/1 | 29.4346 | 28.5555 | 588.9328 | **26.67[a]** |
| 7/26-8/24 | 22.8526 | 20.6895 | 388.5719 | 53.33 |
| 7/18-8/16 | **13.8428[a]** | **10.7964[a]** | **246.3719[a]** | **90[a]** |
| 7/12-8/10 | **14.0029[a]** | **12.9727[a]** | **228.3183[a]** | **96.67[a]** |
| 7/5-8/3 | **10.7706[a]** | **10.5684[a]** | 215.3387 | **98.1[a]** |
| Sub-epidemic model | | | | |
| 9/28-10/27 | **20.5648[a]** | **20.5648[a]** | **488.10[a]** | **26.67[a]** |
| 9/28-10/27 | **20.3471[a]** | **20.3471[a]** | **530.03[a]** | **13.33[a]** |
| 9/21-10/21 | **18.7543[a]** | **18.7543[a]** | **449.43[a]** | 0 |
| 9/14-10/13 | **17.5387[a]** | **17.5387[a]** | **391.48[a]** | **16.67[a]** |
| 9/8-10/8 | **18.5583[a]** | **18.4761[a]** | **419.3323[a]** | 0 |

| | | | | |
|---|---|---|---|---|
| 8/31-9/29 | **24.0052[a]** | **23.723[a]** | **616.2485[a]** | 0 |
| 8/23-9/21 | **22.4065[a]** | **21.7955[a]** | **396.6654[a]** | **6.6[a]** |
| 8/18-9/16 | **18.8891[a]** | **17.5734[a]** | **405.5793[a]** | **40[a]** |
| 8/3-9/1 | **19.5533[a]** | **18.7346[a]** | **415.5[a]** | 23.33 |
| 7/26-8/24 | **9.0521[a]** | **7.7167[a]** | **146.8858[a]** | **100[a]** |
| 7/18-8/16 | 42.8949 | 40.2995 | 442.028 | 26.67 |
| 7/12-8/10 | 32.6091 | 28.1274 | 265.9325 | 50 |
| 7/5-8/3 | 23.8153 | 19.1481 | **160.4378[a]** | 76.67 |

[a]Best performance with regards to the performance metric (row) per forecasting period, i.e. highest prediction interval (PI) coverage and lowest mean interval score (MIS), mean squared error (MSE) and mean absolute error (MAE).

Table S4: Forecasting performance metrics for the country of Mexico as of November 11, 2020.

Higher 95% PI coverage and lower RMSE, MAE and MIS represent better performance.

| Forecast period | RMSE | MAE | MIS | PI |
|---|---|---|---|---|
| GLM | | | | |
| 9/28-10/27 | 105.8058 | 97.4216 | 3313.2 | 0 |
| 9/28-10/27 | 111.0911 | 103.1692 | 3481.3 | 0 |
| 9/21-10/21 | 86.1561 | 82.4714 | 2554.8 | 0 |
| 9/14-10/13 | **61.7137[a]** | **59.1998[a]** | **1642.5[a]** | 13.33 |
| 9/8-10/8 | 56.4818 | 53.3631 | 1261.9 | 30 |
| 8/31-9/29 | 49.8691 | 44.3265 | 1042.8 | 46.67 |
| 8/23-9/21 | **28.141[a]** | **20.9474[a]** | **158.4823[a]** | **80[a]** |
| 8/18-9/16 | **25.894[a]** | **23.1539[a]** | **905.553[a]** | **100[a]** |
| 8/3-9/1 | 158.1838 | 145.7292 | **145.7292[a]** | **97.79[a]** |
| 7/26-8/24 | 172.1921 | 168.4917 | 5254.9 | 0 |
| 7/18-8/16 | 211.4303 | 184.3875 | 5194.8 | 6.67 |
| 7/12-8/10 | 196.9204 | 155.4257 | 3941.1 | 23.33 |
| 7/5-8/3 | 127.4198 | 63.0204 | **331.796[a]** | **96.67[a]** |
| Richards model | | | | |
| 9/28-10/27 | 161.5797 | 156.8476 | 7602.1 | 0 |
| 9/28-10/27 | 151.8023 | 146.9647 | 5424.5 | 0 |
| 9/21-10/21 | 141.3099 | 139.3983 | 6244.4 | 0 |
| 9/14-10/13 | 129.4736 | 128.4193 | 5824.1 | 0 |
| 9/8-10/8 | 125.7501 | 124.2964 | 5118.7 | 0 |
| 8/31-9/29 | 137.8558 | 127.1276 | 4212.4 | 0 |

| | | | | |
|---|---|---|---|---|
| 8/23-9/21 | 118.6926 | 116.2474 | 3977.7 | 0 |
| 8/18-9/16 | 105.069 | 103.5127 | 4023.3 | 0 |
| 8/3-9/1 | 209.7771 | 203.4066 | 6098.5 | 0 |
| 7/26-8/24 | 205.7573 | 202.7084 | 6044.7 | 0 |
| 7/18-8/16 | 226.2236 | 203.7907 | 5423.3 | 10 |
| 7/12-8/10 | 211.1972 | 176.845 | 4275 | *26.67* |
| 7/5-8/3 | 137.025 | 67.7139 | 1278.3 | 73.33 |
| Sub-epidemic model | | | | |
| 9/28-10/27 | **12.8136[a]** | **12.8136[a]** | **583.6016[a]** | **96.67[a]** |
| 9/28-10/27 | **20.3586[a]** | **20.3586[a]** | **595.3998[a]** | **83.33[a]** |
| 9/21-10/21 | **23.9719[a]** | **23.9719[a]** | **540.9622[a]** | **76.67[a]** |
| 9/14-10/13 | 71.6518 | 71.6518 | 1976.6 | **36.67[a]** |
| 9/8-10/8 | **27.0499[a]** | **24.8769[a]** | **530.0952[a]** | **100[a]** |
| 8/31-9/30 | **23.9689[a]** | **23.9689[a]** | **815.282[a]** | **76.67[a]** |
| 8/23-9/21 | 480.1813 | 480.0522 | 17929 | 0 |
| 8/18-9/16 | 222.3915 | 216.1567 | 6843.3 | 0 |
| 8/3-9/1 | **52.669[a]** | **46.9102[a]** | 695.6457 | 60 |
| 7/26-8/24 | **37.45[a]** | **33.7916[a]** | **384.3867[a]** | **86.67[a]** |
| 7/18-8/16 | **94.2513[a]** | **85.8963[a]** | **1954.1[a]** | **23.33[a]** |
| 7/12-8/10 | **64.5419[a]** | **49.9772[a]** | **816.2393[a]** | **56.67[a]** |
| 7/5-8/3 | **67.0493[a]** | **52.9937[a]** | 545.2953 | 80 |

[a]Best performance with regards to the performance metric (row) per forecasting period, i.e. highest prediction interval (PI) coverage and lowest mean interval score (MIS), mean squared error (MSE) and mean absolute error (MAE).
